# ABC_2_-SPH risk score for in-hospital mortality in COVID-19 patients: development, external validation and comparison with other available scores

**DOI:** 10.1101/2021.02.01.21250306

**Authors:** Milena S. Marcolino, Magda C. Pires, Lucas Emanuel F. Ramos, Rafael T. Silva, Luana M. Oliveira, Rafael L.R. Carvalho, Rodolfo L.S. Mourato, Adrián Sánchez-Montalvá, Berta Raventós, Fernando Anschau, José Miguel Chatkin, Matheus C. A. Nogueira, Milton H. Guimarães, Giovanna G. Vietta, Helena Duani, Daniela Ponce, Patricia K. Ziegelmann, Luís C. Castro, Karen B. Ruschel, Christiane C. R. Cimini, Saionara C. Francisco, Maiara A. Floriani, Guilherme F. Nascimento, Bárbara L. Farace, Luanna S. Monteiro, Maira V. R. Souza-Silva, Thais L. S. Sales, Karina Paula M. P. Martins, Israel J. Borges do Nascimento, Tatiani O. Fereguetti, Daniel T. M. O. Ferrara, Fernando A. Botoni, Ana Paula Beck da Silva Etges, Eric Boersma, Carisi A. Polanczyk, Brazilian COVID-19 Registry Investigators

## Abstract

**Objective:** To develop and validate a rapid scoring system at hospital admission for predicting in-hospital mortality in patients hospitalized with coronavirus disease 19 (COVID-19), and to compare this score with other existing ones.

**Design:** Cohort study

**Setting:** The Brazilian COVID-19 Registry has been conducted in 36 Brazilian hospitals in 17 cities. Logistic regression analysis was performed to develop a prediction model for in-hospital mortality, based on the 3978 patients that were admitted between March-July, 2020. The model was then validated in the 1054 patients admitted during August-September, as well as in an external cohort of 474 Spanish patients.

**Participants:** Consecutive symptomatic patients (≥18 years old) with laboratory confirmed COVID-19 admitted to participating hospitals. Patients who were transferred between hospitals and in whom admission data from the first hospital or the last hospital were not available were excluded, as well those who were admitted for other reasons and developed COVID-19 symptoms during their stay.

**Main outcome measures:** In-hospital mortality

**Results:** Median (25th-75th percentile) age of the model-derivation cohort was 60 (48-72) years, 53.8% were men, in-hospital mortality was 20.3%. The validation cohorts had similar age distribution and in-hospital mortality. From 20 potential predictors, seven significant variables were included in the in-hospital mortality risk score: age, blood urea nitrogen, number of comorbidities, C-reactive protein, SpO_2_/FiO_2_ ratio, platelet count and heart rate. The model had high discriminatory value (AUROC 0.844, 95% CI 0.829 to 0.859), which was confirmed in the Brazilian (0.859) and Spanish (0.899) validation cohorts. Our ABC_2_-SPH score showed good calibration in both Brazilian cohorts, but, in the Spanish cohort, mortality was somewhat underestimated in patients with very high (>25%) risk. The ABC_2_-SPH score is implemented in a freely available online risk calculator (https://abc2sph.com/).

**Conclusions:** We designed and validated an easy-to-use rapid scoring system based on characteristics of COVID-19 patients commonly available at hospital presentation, for early stratification for in-hospital mortality risk of patients with COVID-19.

**Summary boxes:** **What is already known on this topic?**

- Rapid scoring systems may be very useful for fast and effective assessment of COVID-19 patients in the emergency department.
- The majority of available scores have high risk of bias and lack benefit to clinical decision making.
- Derivation and validation studies in low- and middle-income countries, including Latin America, are scarce.

**What this study adds**

- ABC_2_-SPH employs seven well defined variables, routinely assessed upon hospital presentation: age, number of comorbidities, blood urea nitrogen, C reactive protein, Spo2/FiO2 ratio, platelets and heart rate.
- This easy-to-use risk score identified four categories at increasing risk of death with a high level of accuracy, and displayed better discrimination ability than other existing scores.
- A free web-based calculator is available and may help healthcare practitioners to estimate the expected risk of mortality for patients at hospital presentation.

## Introduction

Coronavirus disease 2019 (COVID-19), caused by the SARS-CoV-2 virus, is still the main global health, social and economic challenge, overwhelming health care systems in many countries and heavily burdening others, with over 102 million cases and 2.2 million deaths worldwide.^1,2^ While some countries have been declining in new cases, many others have been experiencing a worse surge of the disease than the first wave. Latin America is currently worst-hit region of COVID-19 cases in the world, along with Asia. ^3,4^ Case rates continue to rise, and some hospitals are nearly at their full capacity of intensive care unit beds. The emergence of the new variants of SARS-CoV-2 in England, South Africa and Brazil, with very high viral growth, potentially more transmissible, less detectable with the RT-PCR technique and an unknown response to the available vaccines, is currently a cause of huge concern^5–7^.

Fast and efficient assessment of prognosis of the disease is needed to optimize the allocation of health care and human resources, to empower early identification and intervention of patients at higher risk of poor outcome. A proper assessment tool will guide decision making to develop an appropriate plan of care for each patient^8^. In this context, rapid scoring systems, which combine different variables to estimate the risk of a poor outcome, may be extremely helpful for quick and effective assessment of those patients in the emergency department^9^.

Although different scoring systems have been proposed to assess prognosis in COVID-19 patients, the majority of them lack benefit to clinical decision making, and there is a lack of reliable prognostic prediction models^10,11^. Most scores were developed from small cohorts, at high risk for bias, with selected study samples and relatively few outcome events, without clear details of model derivation and validation, as well as unclear reporting on intended use^12–16^. These issues lead to a high risk of model overfitting, thus their predictive performance when used in clinical practice may be different than that reported.^11,12^ Additionally, clinical characteristics of COVID-19 patients and disease severity vary in different studies in different countries^17^, and external validation was rarely done. Derivation and validation studies in low- and middle-income countries, including Latin America, are scarce^11^.

In this context, our aim was to develop and validate an easy applicable rapid scoring system that employs routinely available clinical and laboratory data at hospital presentation, to predict in-hospital mortality in patients with COVID-19, able to discriminate high vs non-high risk patients. Additionally, we aimed to compare this score with other existing ones.

## Methods

This study is part of the Brazilian COVID-19 Registry, an ongoing multicenter observational study described elsewhere^18^, and a collaboration with Vall d’Hebron University Hospital, in Barcelona, Spain, for independent external validation. The Brazilian COVID-19 Registry is being conducted according to a predefined protocol, in 36 Brazilian hospitals, located in 17 cities, from four Brazilian states. With regards to the type of hospital, 25 are reference centers for COVID-19 treatment and 19 are academic hospitals. Eighteen are public hospitals; seven are private; and eleven are “mixed”, hospitals that provide both public and private services. The median number of hospital beds was 316 (ranging from 60 to 936), and the median number of ICU beds for COVID patients was 22 (ranging from 0 to 105).

Model development, validation and reporting followed guidance from the Transparent Reporting of a Multivariable Prediction Model for Individual Prediction or Diagnosis (TRIPOD) checklist and Prediction model Risk Of Bias ASsessment Tool (PROBAST) (Supplementary Material)^19,20^.

### Study subjects

Consecutive patients with laboratory-confirmed COVID-19 admitted to the participating hospitals from March 1 to September 30, 2020 were enrolled. COVID-19 diagnosis was confirmed according to the World Health Organization guidance^21^. For the purpose of the present study, eligible patients were ≥18 years-old and had completed hospitalization (i.e., discharge or death). Patients who were transferred between hospitals and admission data from the first hospital (as we aimed to develop a score to be used in the first assessment) or the last hospital was not available were excluded, as well those who were admitted for other reasons and developed COVID-19 symptoms during their stay (as their information from the first assessment would be biased, and their profile is different from the other patients) (Figure 1). Those who were admitted for other reasons were excluded. Although patients who were transferred to another hospital where we could not get the final outcome were excluded, a comparison of the clinical characteristics with patients who were included is provided in the Supplemental Material (Table S1).

**Figure 1.**
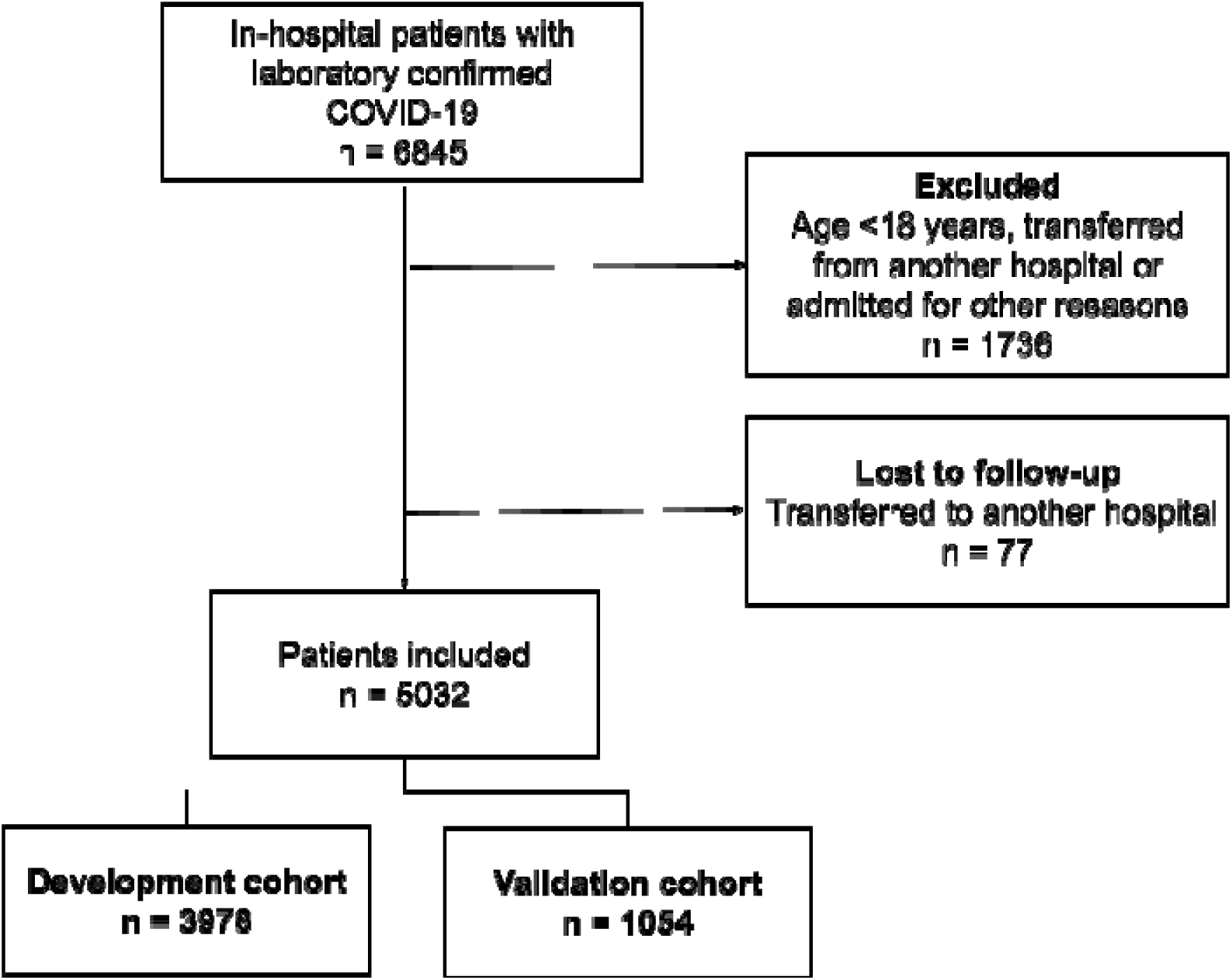
Flowchart of COVID-19 patients included in the study.

### Measurement

Demographic information, clinical characteristics, laboratory and outcome data were collected from the medical records by using a prespecified case report form applying Research Electronic Data Capture (REDCap) tools^22,23^ hosted at the Telehealth Center, University Hospital, *Universidade Federal de Minas Gerais*. Data were collected by trained hospital staff or interns. A detailed data management plan (DMP) was developed and provided to all participating centers. An online DMP training was mandatory before local research personnel were allowed to start collecting study data^24^.

### Data quality assessment

We undertook comprehensive data quality checks to ensure high quality. A code was developed in R software to identify values likely related to data entry errors for vital signs and laboratory variables, based on expert-guided rules. Data were sent to each center for checking and correction. Transfers from one participant hospital to another were merged and considered as a single visit.

### Potential predictors for in-hospital mortality

All variables used to calculate the risk score were obtained at hospital admission. A set of potential predictor variables for in-hospital mortality was selected a priori, as recommended^19^, taking into account the evidence in literature of association with worse prognosis in patients with COVID-19 or pneumonia, and availability of predictor measurement at the time the model would be used, i.e., hospital admission. We considered predictors that would be available in routine practice in most emergency departments worldwide. It included patient demographic characteristics, pre-existing comorbid medical conditions, home medications, clinical assessment at admission and laboratory data ^12^. All laboratory tests were performed at the discretion of the treating physician. Imaging test results were not included, as X-ray and CT scan are not always performed at patient admission and their interpretation involve subjective judgement.

Candidate predictor variables which were not available for at least two thirds of patients within the derivation cohort (more than one third of missing data) were excluded.

### Data analysis

Continuous variables were summarized using medians and interquartile ranges (IQR), whereas we used counts and percentages for categorical variables. We reported 95% confidence intervals, and for all two-tailed-tests performed, a p-value less than 0.05 was considered statistically significant. Statistical analysis was performed with R software (version 4.0.2) with the mgcv, finalfit, mice, glmnet, pROC, rms, rmda, and psfmi packages. Details about how missing data were handled, as well as model-building and model-validation procedures, are described below.

### Missing data

Considering missing at random after analyzing missing data patterns, multiple imputation with chained equations (MICE) was used to handle missing values only on candidate variables (outcomes were not imputed). Outcome variable was considered as a predictor only in the derivation dataset. We used predictive mean matching (PMM) method for continuous predictors and polytomous regression for categorical variables (two or more unordered levels). The results of 10 imputed datasets, each with 10 iterations, were combined following Rubin’s rules^25^.

### Development of the risk score model

Patients who were admitted before July 31 were included in the development cohort. First, we conducted predictor selection based on clinical reasoning and literature review before modeling. Second, generalized additive models (GAM) were used to examine the relationships between in-hospital mortality and continuous (through penalized thin plate splines) and categorical (as linear components) predictors. During this stage, variable selection was based on D1- (multivariate Wald test) and D2-statistic (pools test statistics from the repeated analyses).

Third, for an easier application of the risk score model at bedside, continuous variables were categorized based on widely accepted cut points, current evidence and/or categories defined in stablished rapid scoring systems from pneumonia and sepsis.

Lastly, we used least absolute shrinkage and selection operator (LASSO) logistic regression to derive the mortality score by scaling the (L1 penalized) shrunk coefficients. The penalty parameter λ in LASSO was chosen using 10-fold cross-validation methods based on mean squared error criterion.

Risk groups were proposed based on predicted probabilities: low risk (< 6.0%), intermediate risk (6.0 – 14.9%), high risk (15.0 – 49.9%), and very high risk (≥ 50.0%).

### External validation

We performed an external (temporal) validation analysis using patients who were admitted from August 1 to September 30, 2020. The same investigators collected those data, and missing data were handle as described above.

Independent external validation was also performed in a cohort of patients from Vall d’Hebron University Hospital (Vall d’Hebron COVID-19 Prospective Cohort Study), a 1100-bed public tertiary care hospital with the capacity for more than 60 ICU beds, in Barcelona, Spain^26^, part of the public hospital network of the Catalan Health System. Inclusion and exclusion criteria were the same as the beforementioned ones. All patients included were followed for at least 28 days.

### Performance measures

We evaluated overall performance using Brier score^27^. Calibration was assessed graphically by plotting the predicted mortality probabilities against the observed mortality, testing intercept equals 0 and slope equals 1. The area under the curve for receiver operating characteristic (AUROC) described model’s discrimination, i.e, its ability to predict higher risks for individuals who died than for those who were discharged. Confidence intervals (95% CI) for AUROC were obtained through 2000 bootstrap samples. We also calculated positive and negative predictive values of the derived risk groups.

### Model comparisons

The developed model was compared within the validation cohort with existing rapid scores systems for in-hospital mortality in COVID-19 patients. These scores were identified through a literature search of Medline, medRxiv and BioRxiv, with no language or date restrictions, using the search terms “COVID-19,” “COVID”, “SARS-CoV-2,” “coronavirus” combined with “score” and “mortality”. The last search was performed on November 19, 2020. Two authors independently performed article selection and data extraction. Additionally, we also included established scores for pneumonia and sepsis^28–32^.

From the set of identified scores, we selected those which with predictors were available within the database and had accessible methods for calculation. Model comparisons were performed using AUROC and decision curve analysis, which describes clinical utility across a range of threshold risks, i.e, the relative value of benefits (if a true positive case is treated) and harms (if a false positive case is treated).

### ABC_2_-SPH risk score calculator

Risk score calculator was developed in Javascipt, using the Svelte framework while the website was developed in R language (blogdown package).

### Ethics

The study protocol conforms to the ethical guidelines of the Declaration of Helsinki. It was approved by the Brazilian National Commission for Research Ethics (CAAE 30350820.5.1001.0008) Individual informed consent was waived due to the severity of the situation and the use of deidentified data, based on medical chart review only. For the independent external validation cohort, it was approved by the and Vall d’Hebron University Hospital Research Ethics Committee (PR(AG)183/2020). The institutional review board granted an informed consent waiver if patients were unable to give oral consent.

### Patient and public involvement

This was an urgent public health research study in response to a Public Health Emergency of International Concern. Patients or the public were not involved in the design, conduct, interpretation or presentation of results of this research.

## Results

The derivation cohort comprehended data from 3978 patients, from 267 cities of 13 states in Brazil (Figure 2). The median age was 60 [IQR, 48-72] years, 2138 (53.8%) were male, 2789 (70.1%) had at least one comorbidity and 806 (20.3%) died during hospitalization. The median follow-up time was 7 (4-14) days. Table 1 shows demographic, clinical characteristics and laboratory findings for the derivation and validation datasets.

**Figure 2.**
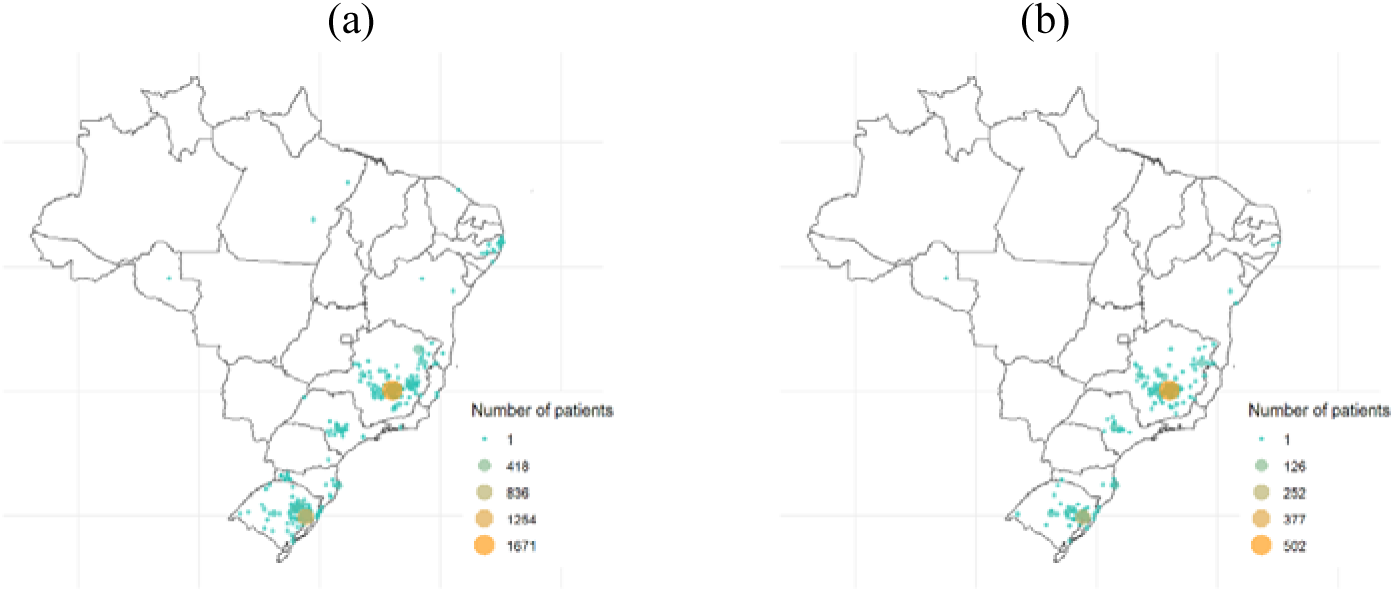
City of residence of patients within (a) development and (b) validation cohorts.

**Table 1.** Demographic and clinical characteristics for derivation and validation cohorts of patients admitted to hospital with COVID-19.

| Characteristics | Derivation cohort (n=3,978) |  | Brazilian validation cohort (n=1,054) |  | Spanish validation cohort (n=474) |  |
| --- | --- | --- | --- | --- | --- | --- |
|  | Frequency (%)<br>or<br>median (IQR) | Non missing<br>cases (%) | Frequency (%) or<br>median (IQR) | Non missing<br>cases (%) | Frequency (%) or<br>median (IQR) | Non missing cases<br>(%) |
| In hospital mortality | 806 (20.3%) | 3,978 (100%) | 208 (19.7%) | 1,054 (100%) | 82 (17.3%) | 474 (100%) |
| Age (years) | 60.0 (48.0,<br>72.0) | 3,978 (100%) | 62.0 (48.2, 73.0) | 1,054 (100%) | 59.5 (49.0, 71.0) | 474 (100%) |
| Sex at birth |  | 3,976 (99.9%) |  | 1,054 (100%) |  | 474 (100%) |
| Male | 2,138 (53.8%) |  | 582 (55.2%) |  | 276 (58.2%) |  |
| **Comorbidities** |  |  |  |  |  |  |
| Hypertension | 2,147 (54.0%) | 3,978 (100%) | 563 (53.4%) | 1,054 (100%) | 193 (40.7%) | 474 (100%) |
| Coronary artery disease | 215 (5.4%) | 3,978 (100%) | 60 (5.7%) | 1,054 (100%) | 32 (6.8%) | 474 (100%) |
| Heart failure | 269 (6.8%) | 3,978 (100%) | 58 (5.5%) | 1,054 (100%) | 23 (4.9%) | 474 (100%) |
| Atrial fibrillation or flutter | 139 (3.5%) | 3,978 (100%) | 27 (2.6%) | 1,054 (100%) | 44 (9.3%) | 474 (100%) |
| Stroke | 146 (3.7%) | 3,978 (100%) | 43 (4.1%) | 1,054 (100%) | 18 (3.8%) | 474 (100%) |
| COPD | 253 (6.4%) | 3,978 (100%) | 60 (5.7%) | 1,054 (100%) | 24 (5.1%) | 474 (100%) |
| Diabetes mellitus | 1,151 (28.9%) | 3,978 (100%) | 297 (28.2%) | 1,054 (100%) | 83 (17.5%) | 474 (100%) |
| Obesity (BMI>30kg/m<sup>2</sup>) | 696 (17.5%) | 3,978 (100%) | 181 (17.2%) | 1,054 (100%) | 112 (23.6%) | 474 (100%) |
| Cirrhosis | 25 (0.6%) | 3,978 (100%) | 9 (0.9%) | 1,054 (100%) | 3 (0.6%) | 474 (100%) |
| Cancer | 194 (4.9%) | 3,978 (100%) | 65 (6.2%) | 1,054 (100%) | 19 (4.0%) | 474 (100%) |
| Number of comorbidities |  | 3,978 (100%) |  | 1,054 (100%) |  | 474 (100%) |
| 0 | 1,189 (29.9%) |  | 309 (29.3%) |  | 195 (41.1%) |  |
| 1 | 1,173 (29.5%) |  | 328 (31.1%) |  | 111 (23.4%) |  |
| 2 | 1,013 (25.5%) |  | 269 (25.5%) |  | 95 (20.0%) |  |
| 3 | 429 (10.8%) |  | 106 (10.1%) |  | 53 (11.2%) |  |
| 4 | 131 (3.3%) |  | 33 (3.1%) |  | 14 (3.0%) |  |
| ≥ 5 | 43 (1.1%) |  | 9 (0.9%) |  | 6 (1.2%) |  |
| **Clinical assessment at admission** |  |  |  |  |  |  |
| SF ratio | 428.6 (332.1, 452.4) | 3,845 (96.7%) | 433.3 (339.3, 452.4) | 1,034 (98.1%) | 459.5 (428.6, 471.4) | 474 (100%) |
| Respiratory rate (irpm) | 20.0 (18.0, 24.0) | 3,236 (81.3%) | 20.0 (18.0, 24.0) | 870 (82.5%) | 20.0 (18.0, 28.0) | 452 (95.3%) |
| Heart rate (bpm) | 88.0 (78.0, 100.0) | 3,787 (95.2%) | 88.0 (77.0, 100.0) | 1,020 (96.8%) | 95.0 (82.0, 108.0) | 474 (100%) |
| Glasgow coma score | 15.0 (15.0, 15.0) | 3,695 (92.9%) | 15.0 (15.0, 15.0) | 982 (93.2%) | 15.0 (15.0, 15.0) | 466 (98.3%) |
| Systolic blood pressure |  | 3,762 (94.6%) |  | 1,014 (96.2%) |  | 471 (99.4%) |
| ≥ 90 (mm Hg) | 3,076 (81.8%) |  | 825 (81.4%) |  | 466 (98.9%) |  |
| < 90 (mm Hg) | 510 (13.6%) |  | 146 (14.4%) |  | 5 (1.1%) |  |
| Inotrope requirement | 176 (4.7%) |  | 43 (4.2%) |  | 0 |  |
| Diastolic blood pressure |  | 3,776 (94.9%) |  | 1,022 (97.0%) |  | 471 (99.4%) |
| > 60 (mm Hg) | 3,541 (93.8%) |  | 962 (94.1%) |  | 405 (86.0%) |  |
| ≤ 60 (mm Hg) | 59 (1.6%) |  | 17 (1.7%) |  | 66 (14.0%) |  |
| Inotrope requirement | 176 (4.7%) |  | 43 (4.2%) |  | 0 |  |
| **Laboratory parameters** |  |  |  |  |  |  |
| Hemoglobin (g/L) | 13.3 (12.1, 14.4) | 3,871 (97.3%) | 13.3 (11.9, 14.5) | 1,021 (96.9%) | 13.4 (12.2, 14.7) | 474 (100%) |
| Platelet count (10<sup>9</sup>/L) | 196.0 (154.0, 257.0) | 3,824 (96.1%) | 203.0 (154.0, 260.2) | 1,016 (96.4%) | 197.5 (155.3, 257.0) | 474 (100%) |
| NLR | 4.7 (2.8, 7.8) | 3,759 (94.5%) | 4.9 (3.0, 8.4) | 989 (93.8%) |  |  |
| Lactate (mmol/L) | 1.4 (1.1, 1.9) | 2,742 (68.9%) | 1.5 (1.2, 2.1) | 720 (68.3%) |  |  |
| C-reactive protein (mg/L) | 77.0 (38.0, 143.0) | 3,487 (87.7%) | 74.1 (33.8, 143.0) | 881 (83.6%) | 102.4 (43.9, 189.3) | 474 (100%) |
| BUN (mg/dL) | 16.3 (11.5, 24.3) | 3,636 (91.4%) | 17.3 (12.9, 25.2) | 942 (89.4%) | 15.9 (11.7, 24.3) | 474 (100%) |
| Creatinine (mg/dL) | 0.9 (0.8, 1.2) | 3,765 (94.6%) | 1.0 (0.8, 1.3) | 967 (91.7%) | 0.8 (0.7, 1.1) | 474 (100%) |
| Sodium (mmol/L) | 137.0 (135.0, 140.0) | 3,550 (89.2%) | 137.0 (134.3, 140.0) | 930 (88.2%) | 136.0 (134.2, 138.0) | 474 (100%) |
| Bicarbonate (mEq/L) | 23.0 (21.0, 25.0) | 3,222 (81.0%) | 23.0 (20.6, 25.0) | 807 (76.6%) | NA | NA |
| pH | 7.4 (7.4, 7.5) | 3,232 (81.2%) | 7.4 (7.4, 7.5) | 808 (76.7%) | NA | NA |
| pO2 (mmHg) | 75.0 (63.0, 96.0) | 3,183 (80.0%) | 73.4 (63.0, 94.6) | 800 (75.9%) | NA | NA |
| pCO2 (mmHg) | 35.0 (31.3, 39.0) | 3,194 (80.3%) | 34.0 (30.0, 38.0) | 801 (76.0%) | NA | NA |

BMI: body mass index; BUN: blood urea nitrogen; COPD: chronic obstructive pulmonary disease; NA: not available; NLR: neutrophils-to-lymphocytes ratio; SF ratio: SpO<sub>2</sub>/FiO<sub>2</sub> ratio

### Development of the risk score model

Thirty-six potential predictor variables were identified (Table S2). Number of comorbidities was created as a composite of ten individual comorbidities shown to have prognostic impact in COVID-9 (hypertension, diabetes mellitus, obesity, coronary artery disease, heart failure, atrial fibrillation or flutter, cirrhosis, cancer and previous stroke)^33,34^, as in other scores^35,36^. Twelve variables were excluded due to the excessive number of missing values, two for high collinearity, and one was not recorded within database. Besides that, inotrope use was combined with blood pressure. Therefore, 20 variables were tested.

Through generalized additive model (GAM), a combination of seven variables was selected as the best predictor of in hospital mortality (Table S3). For an easier application to the risk score model at bedside, continuous selected predictors were categorized for LASSO logistic regression. All categories were defined a priori, as recommended,^20^ based on widely accepted cut points, current evidence and/or categories defined in stablished rapid scoring systems from pneumonia and sepsis, as follows: advanced age (60-69.9, 70-79.9 and ≥ 80 years), Spo2/Fio2 (SF) ratio (≤ 150.0, 150.1 – 235.0, 235.1 – 315.0, > 315.0), platelet count (<100×10^9^/L, 100-150×10^9^/L, > 150×10^9^/L), C-reactive protein (≥100mg/L), blood urea nitrogen (BUN) (≥42mg/dL), heart rate (≤ 90, 91-130, ≥ 131 bpm).

All variables were statistically significant predictors for in hospital mortality (Table S4 and Figure S1). Shrunk coefficients were scaled to provide a prognostic index and we denoted it as the ABC_2_-SPH risk score (Table 2). The sum of the prediction scores ranges between 0 and 20, with a high score indicating higher risk of in-hospital mortality.

**Table 2.** ABC_2_-SPH Score for in-hospital mortality in patients with COVID-19.

|  | Variable | ABC<sub>2</sub>-SPH score |
| --- | --- | --- |
| **A** | **Age (years)** |  |
|  | < 60 | 0 |
|  | 60 - 69 | 1 |
|  | 70 - 79 | 3 |
|  | ≥ 80 | 5 |
| **B** | **Blood urea nitrogen (mg/dL)\*** |  |
|  | < 42 | 0 |
|  | ≥ 42 | 3 |
| **C<sub>2</sub>** | **Comorbidities** |  |
|  | 0 – 1 | 0 |
|  | ≥ 2 | 1 |
|  | **C reactive protein (mg/L)** |  |
|  | < 100 | 0 |
|  | ≥ 100 | 1 |
| **S** | **SF ratio (%)** |  |
|  | > 315.0 | 0 |
|  | 235.1 – 315.0 | 1 |
|  | 150.1 – 235.0 | 3 |
|  | ≤ 150.0 | 6 |
| **P** | **Platelet count (x10<sup>9</sup>/L)** |  |
|  | > 150 | 0 |
|  | 100 -150 | 1 |
|  | < 100 | 2 |
| **H** | **Heart rate (bpm)** |  |
|  | ≤ 90 | 0 |
|  | 91 – 130 | 1 |
|  | ≥ 131 | 2 |

\* When converted to urea, the cut-off is 90 mg/dL.

Risk groups were proposed based on predicted probabilities (Table 3): low risk (0-1 score, observed in hospital mortality 2.0%), intermediate risk (2-4 score, 11.4%), high risk (5-8 score, 32.0%), and very high risk (≥ 9 score, 69.4%). Subject-specific risks can be assessed using the developed ABC_2_-SPH risk score Web-based calculator (https://abc2sph.com/), freely available to the public.

**Table 3.** Predicted mortality and mortality rates for ABC_2_-SPH Score risk groups.

| Risk Group | Predicted mortality | Derivation cohort |  | Validation cohort |  |
| --- | --- | --- | --- | --- | --- |
|  |  | No of patients | No of deaths (%) | No of patients | No of deaths (%) |
| Low (0-1) | < 6% | 1133 | 23 (2.0%) | 290 | 1 (0.3%) |
| Intermediate (2-4) | 6 - 14.9% | 1470 | 168 (11.4%) | 394 | 47 (11.9%) |
| High (5-8) | 15 - 49.9% | 907 | 290 (32.0%) | 252 | 73 (29.0%) |
| Very high (≥9) | ≥ 50% | 468 | 325 (69.4%) | 118 | 87 (73.7%) |
| Overall | - | 3978 | 806 (20.3%) | 1054 | 208 (19.7%) |

As well as GAM and LASSO, ABC_2_-SPH risk score showed good overall performance (Brier score: 0.114) and good discrimination (AUROC equal 0.842; 95% CI 0.840–0.843) within the derivation cohort (Table 4).

**Table 4.**
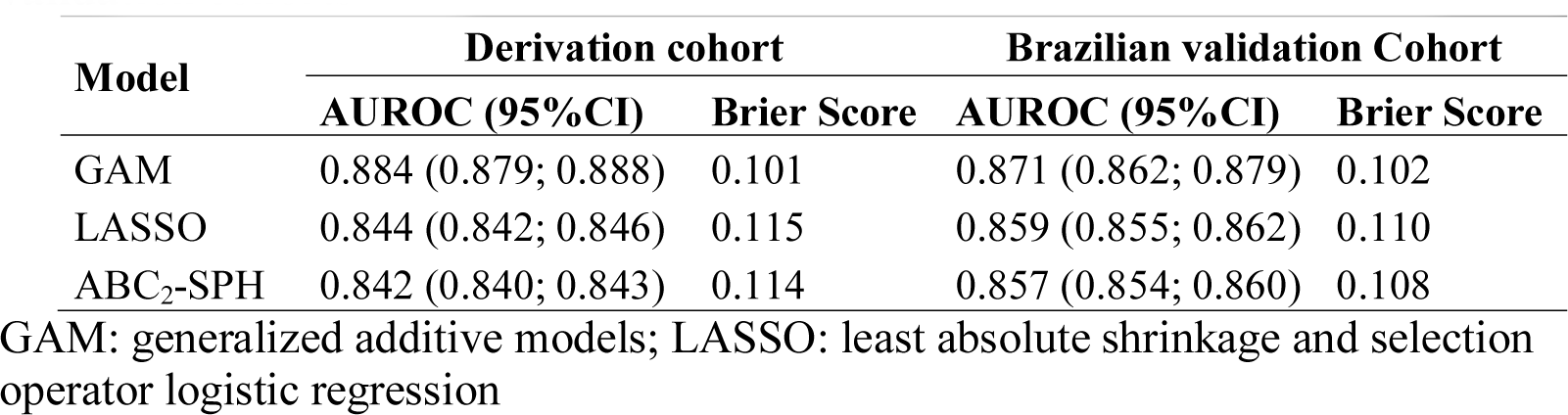
Discrimination and model overall performance in derivation and validation cohorts.

| Model | Derivation cohort |  | Brazilian validation Cohort |  |
| --- | --- | --- | --- | --- |
|  | AUROC (95%CI) | Brier Score | AUROC (95%CI) | Brier Score |
| GAM | 0.884 (0.879; 0.888) | 0.101 | 0.871 (0.862; 0.879) | 0.102 |
| LASSO | 0.844 (0.842; 0.846) | 0.115 | 0.859 (0.855; 0.862) | 0.110 |
| ABC<sub>2</sub>-SPH | 0.842 (0.840; 0.843) | 0.114 | 0.857 (0.854; 0.860) | 0.108 |

GAM: generalized additive models; LASSO: least absolute shrinkage and selection operator logistic regression

### External validation – Brazilian cohort

A total of 1054 patients were included in the validation cohort. The median age was 62 (interquartile range 48-73) years, 582 (55.2%) were male and 745 (70.7%) had at least one comorbidity. The median follow-up time was 7 (4-13) days. Two hundred and eight patients (19.7%) died during hospitalization. The distribution of patients across range ABC_2_-SPH Score in derivation and validation cohorts are presented in Figure 3.

**Figure 3.**
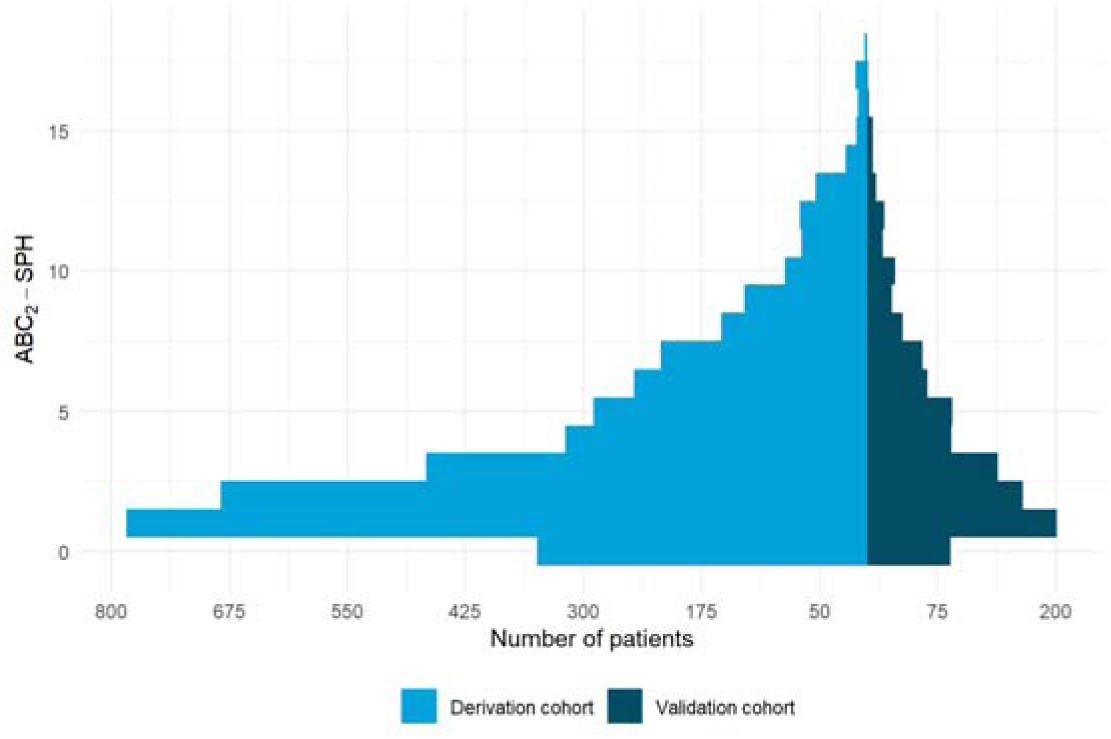
ABC_2_-SPH Score in derivation and validation cohorts.

We observed good discrimination (AUROC equal 0.859; 95% CI 0.833 to 0.885; Figure 4), overall performance (Brier = 0.108) and calibration (slope = 1.138, intercept = 0.114, p-value = 0.184; Figure S2a) of the ABC_2_-SPH risk score under the validation cohort (Figure 4). The good performance is also demonstrated in sensitivity analyses using complete case data (Table S5).

**Figure 4.**
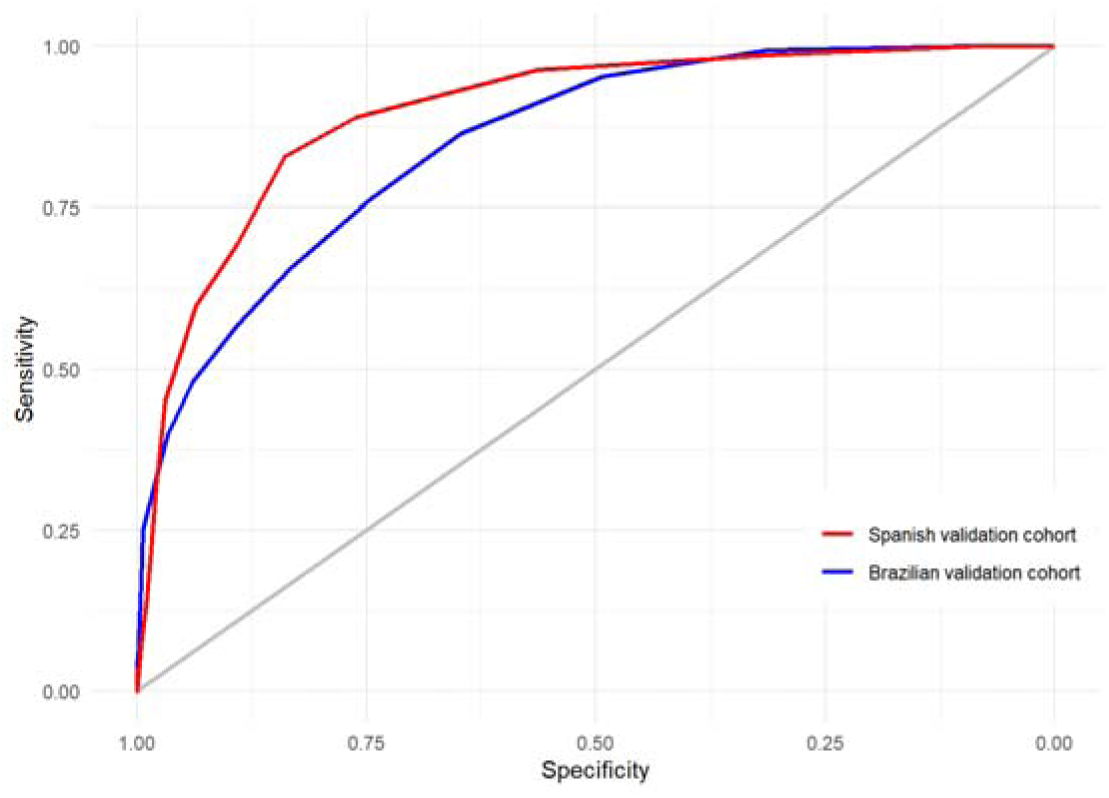
Discrimination of ABC_2_-SPH Score in external validation cohorts.

Low, intermediate and high-risk groups showed good negative predictive values (99.7%, 88.1% and 71.0%, respectively). A positive predictive value of 73.7% was observed in patients classified as at very high mortality risk.

### External validation – Spanish cohort

A second external (geographic) validation was performed within a Spanish cohort with 474 patients and 82 (17,3%) in hospital mortality. The demographic and clinical characteristics at admission are listed in Table 1. The median follow-up time was 21 (IQR, 7-40) days. Only complete cases were included.

ABC_2_-SPH Score showed high discrimination (AUROC= 0.899, 95% CI 0.864 to 0.934; Figure 4), good overall performance (Brier = 0.093), but an underestimation of true mortality risk in patients with a predicted probability above 25% (intercept = 0.729, slope = 1.519, p-value = 0.001; Figure S2b).

### Literature review

The literature search identified 39 scores to predict mortality in COVID-19 patients (Table 5). Most of them were still preprints (28%), in 36% the derivation cohort was from China, 21% from the United States and none from South America. Multivariate logistic regression and LASSO regression were used in 16 and 10 studies, respectively, artificial intelligence techniques in seven studies and Cox regression analysis in 3 studies. Two scores were developed by consensus. The population of the development cohort was composed by adults-only in 51.3% of the studies, the age range was not clear in 41.4% and elderly patients in one of them. Thirteen studies developed points-based scores, three were published as nomograms and all the other ones required formulas for calculation.

**Table 5.**
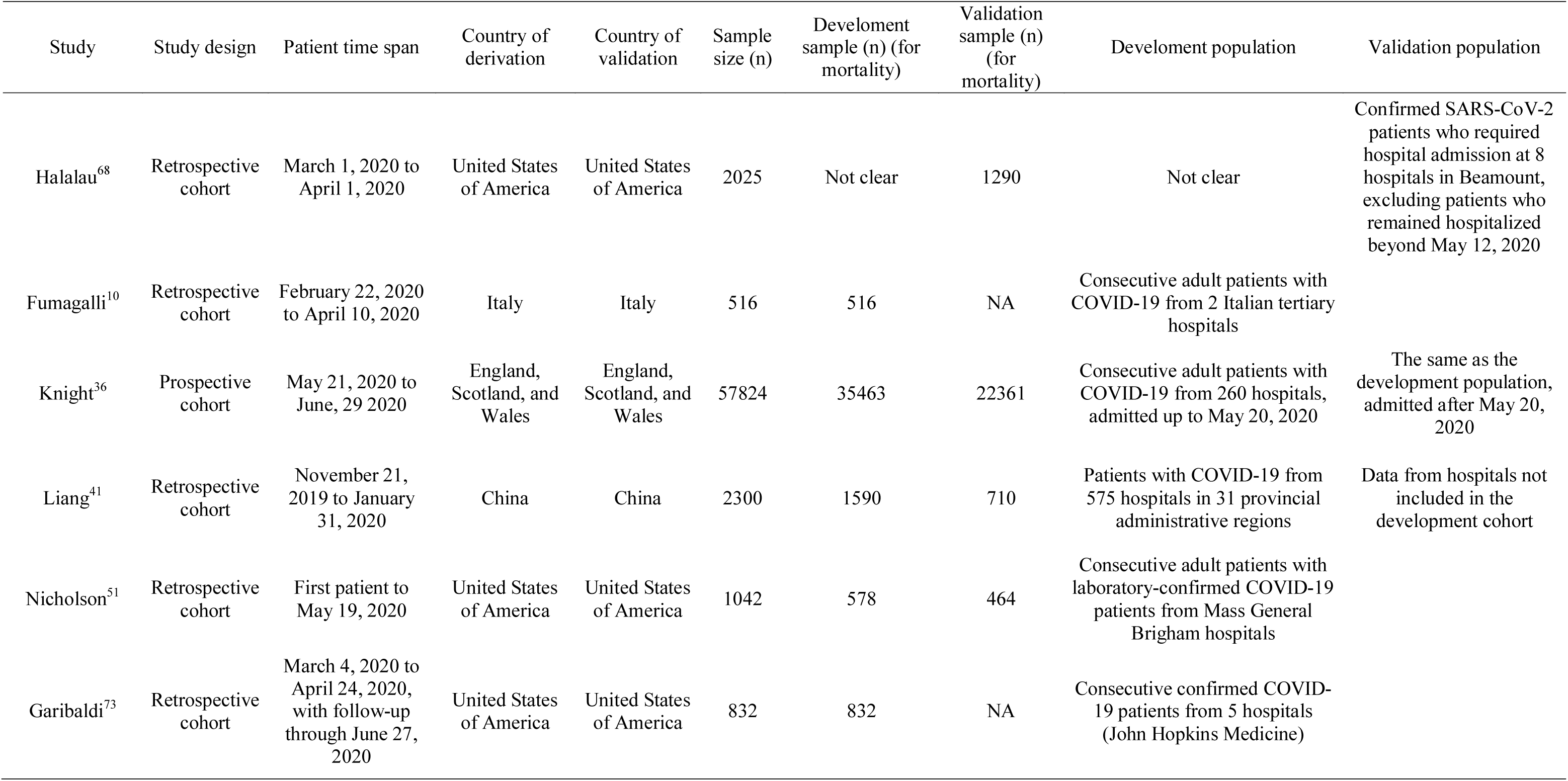
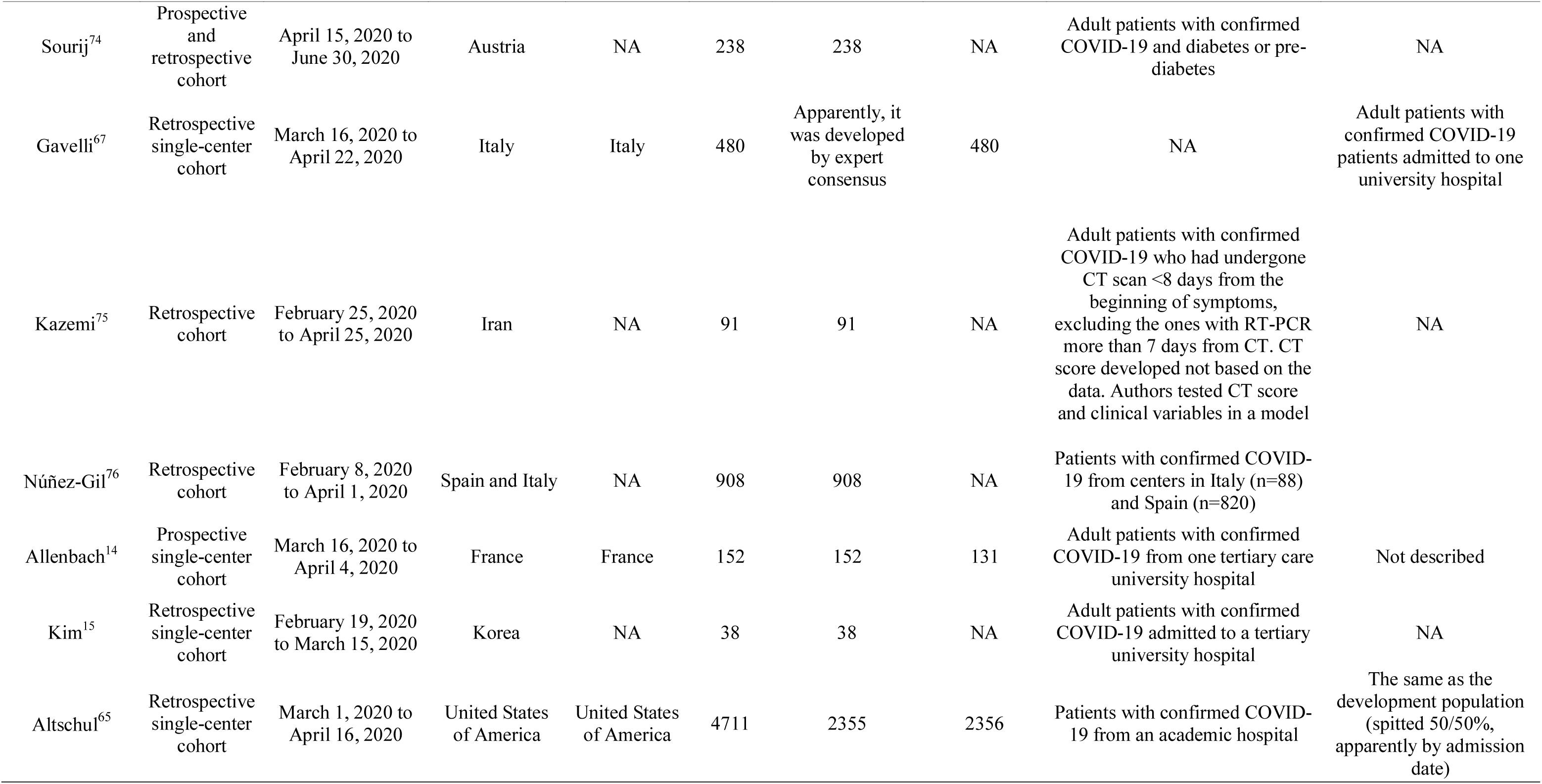
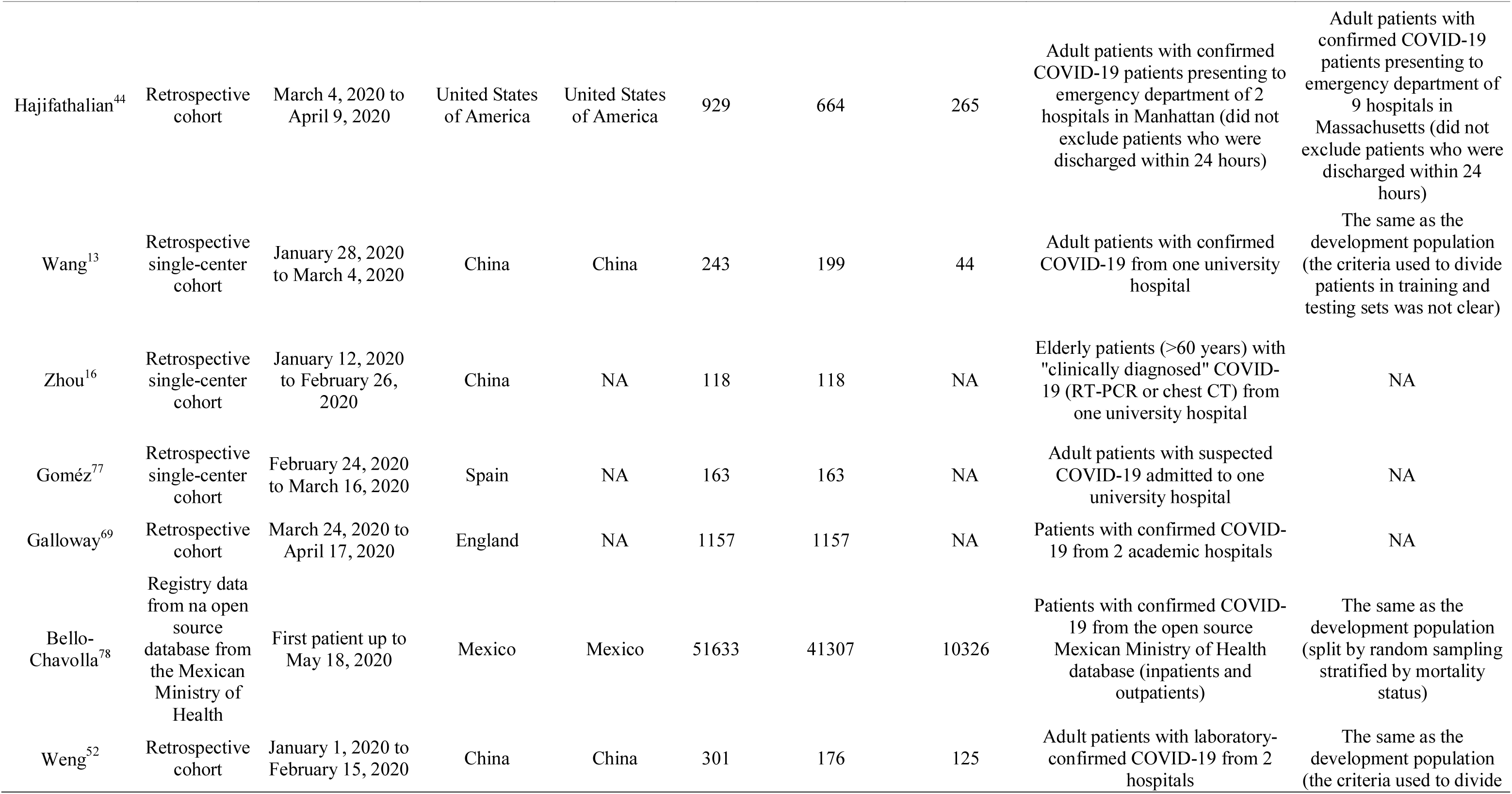
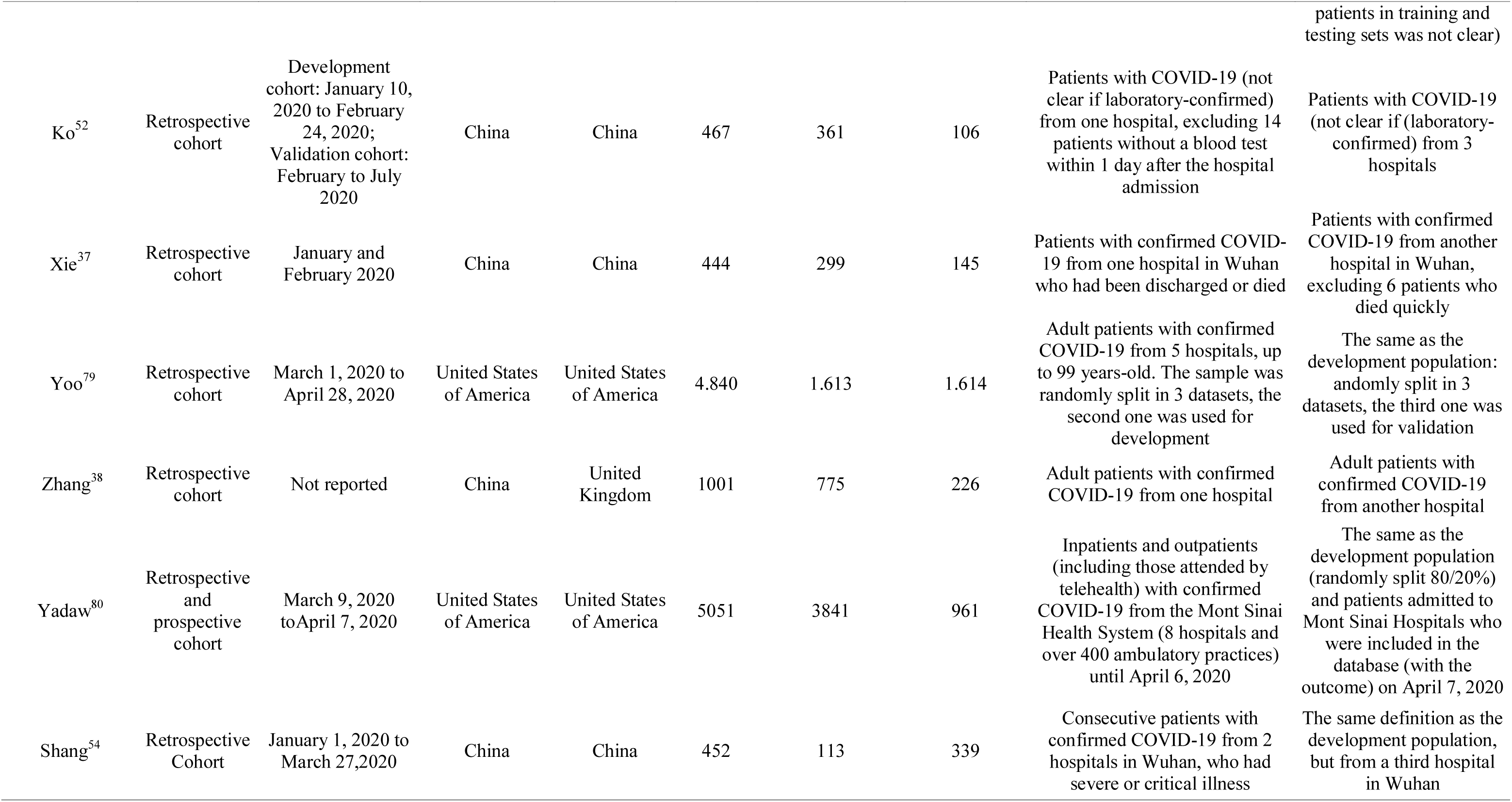
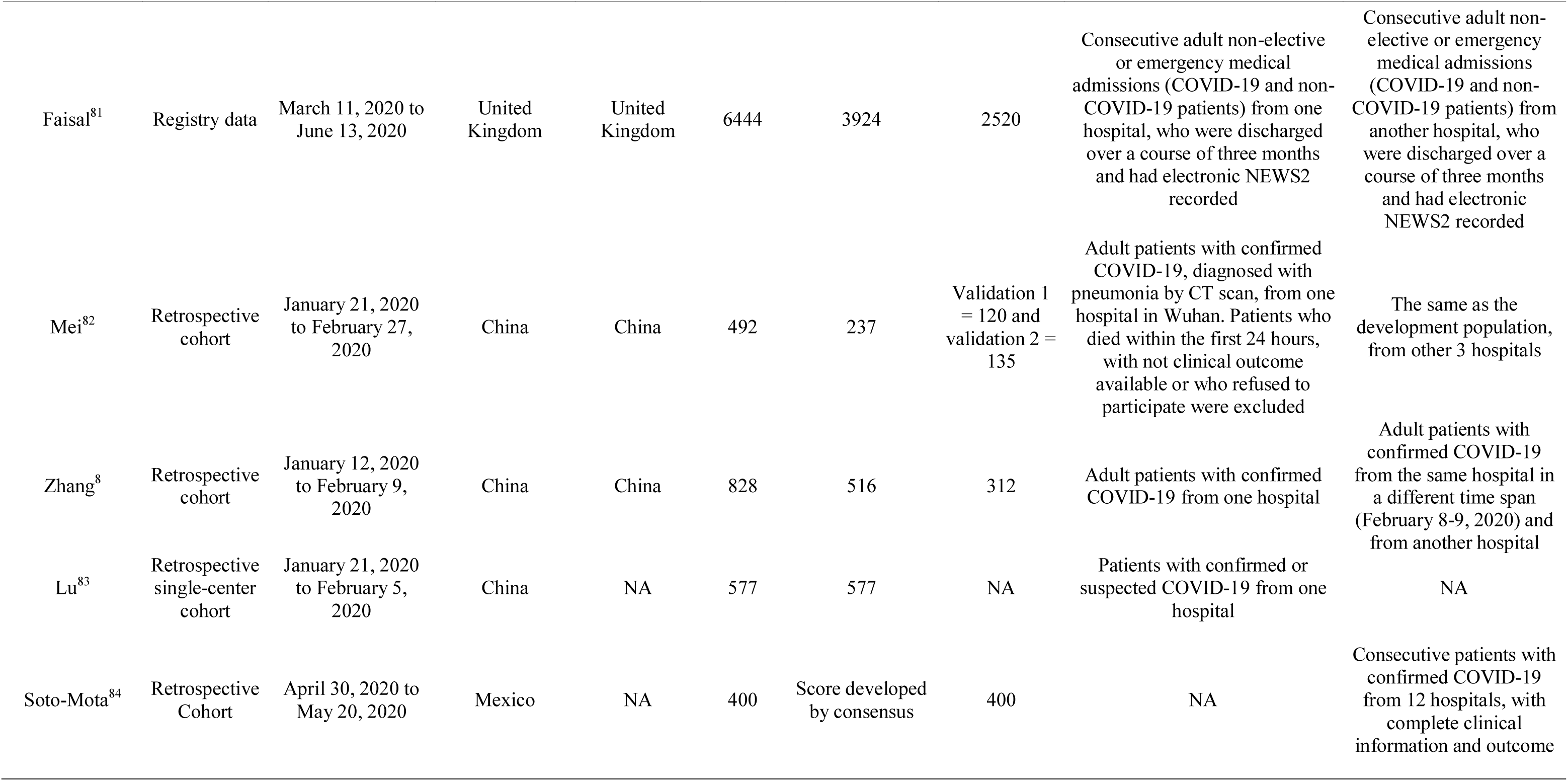
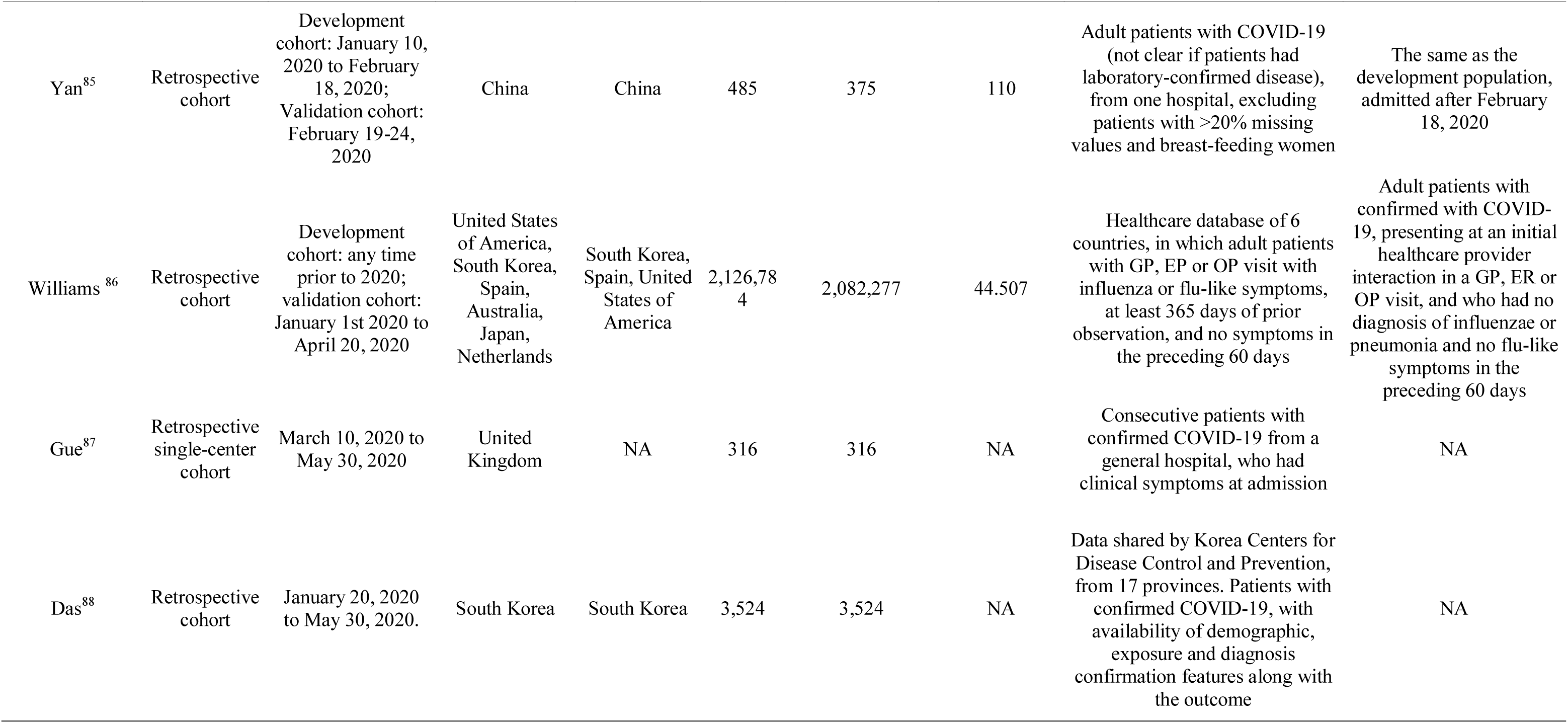
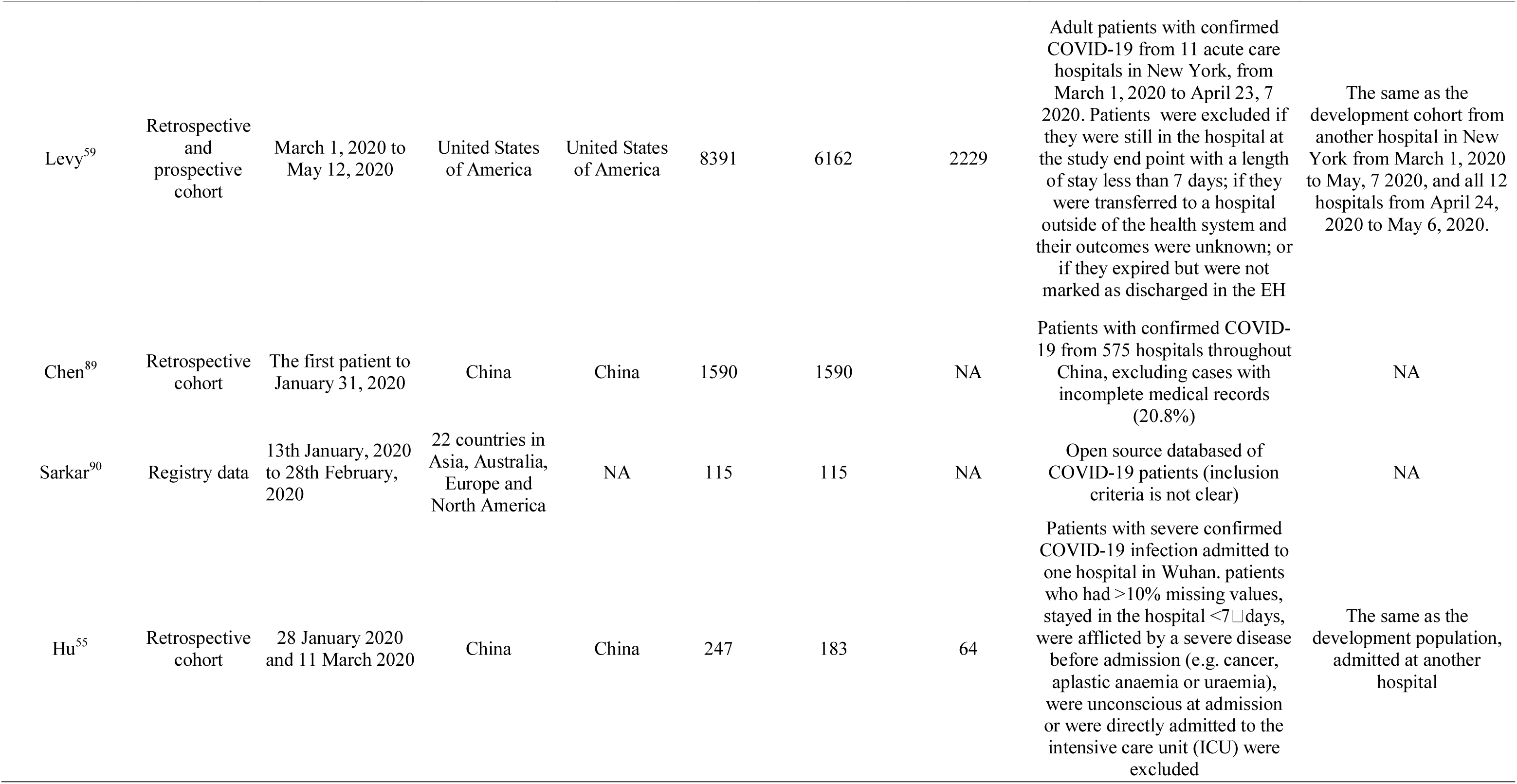
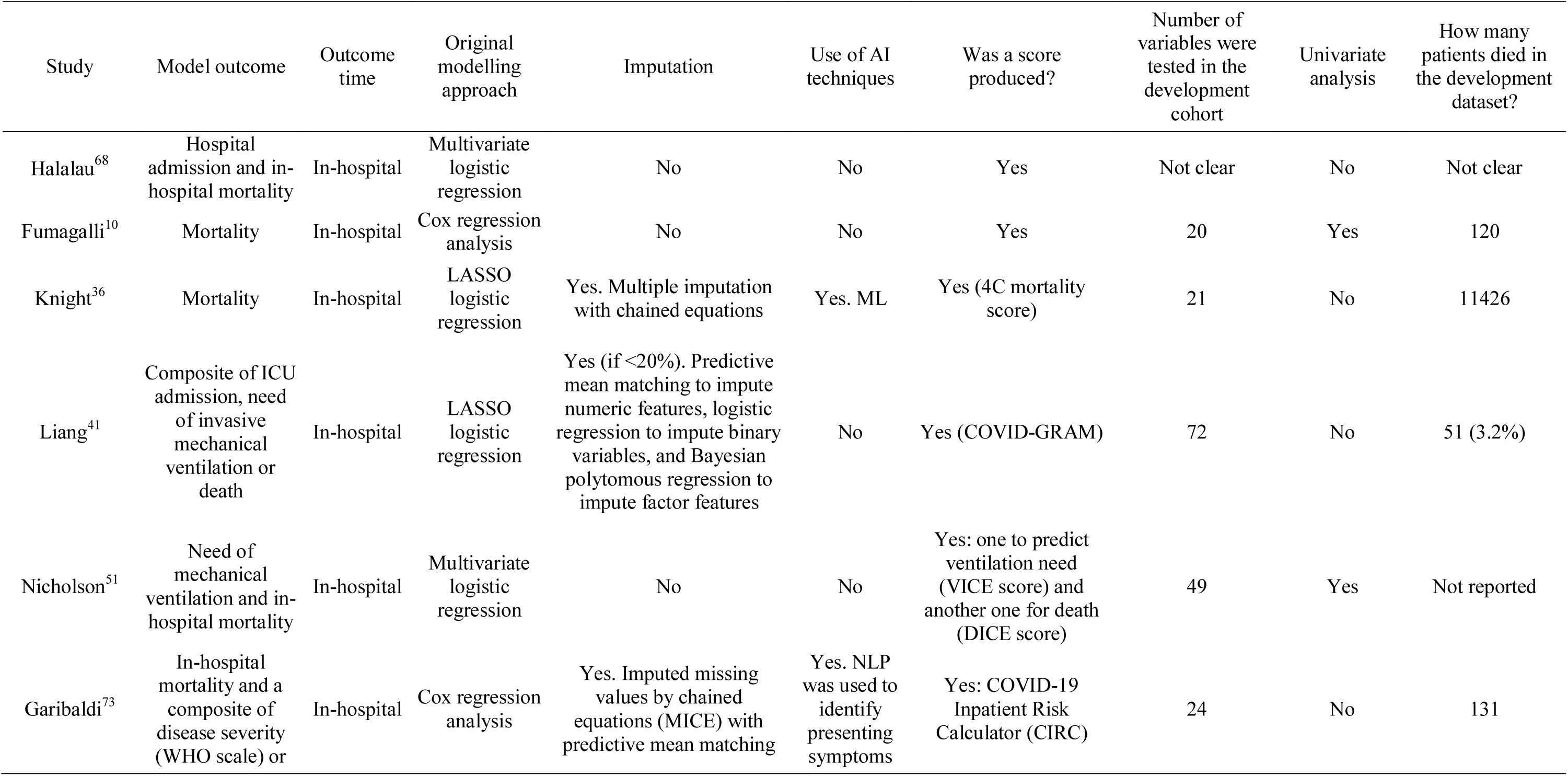
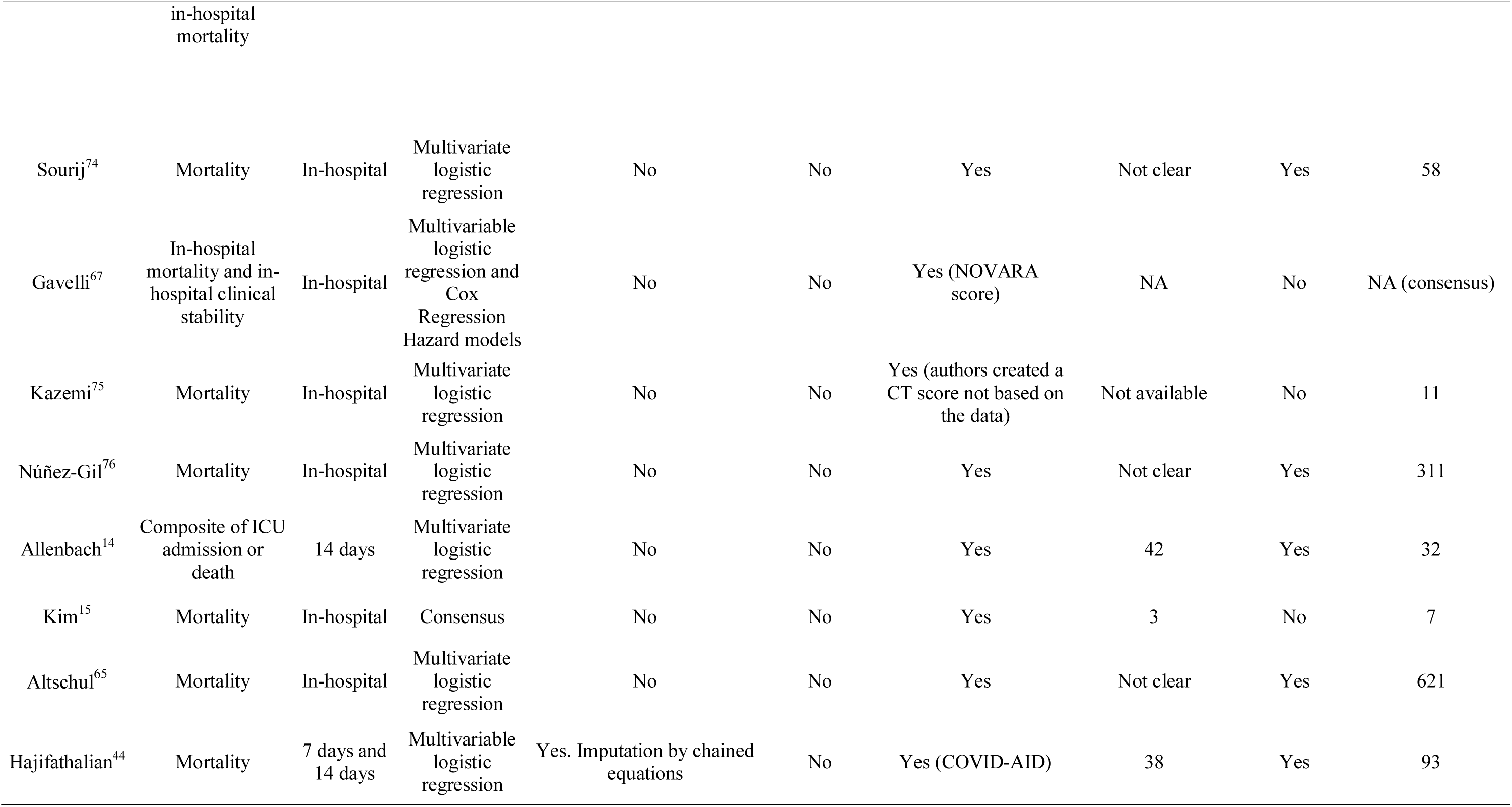
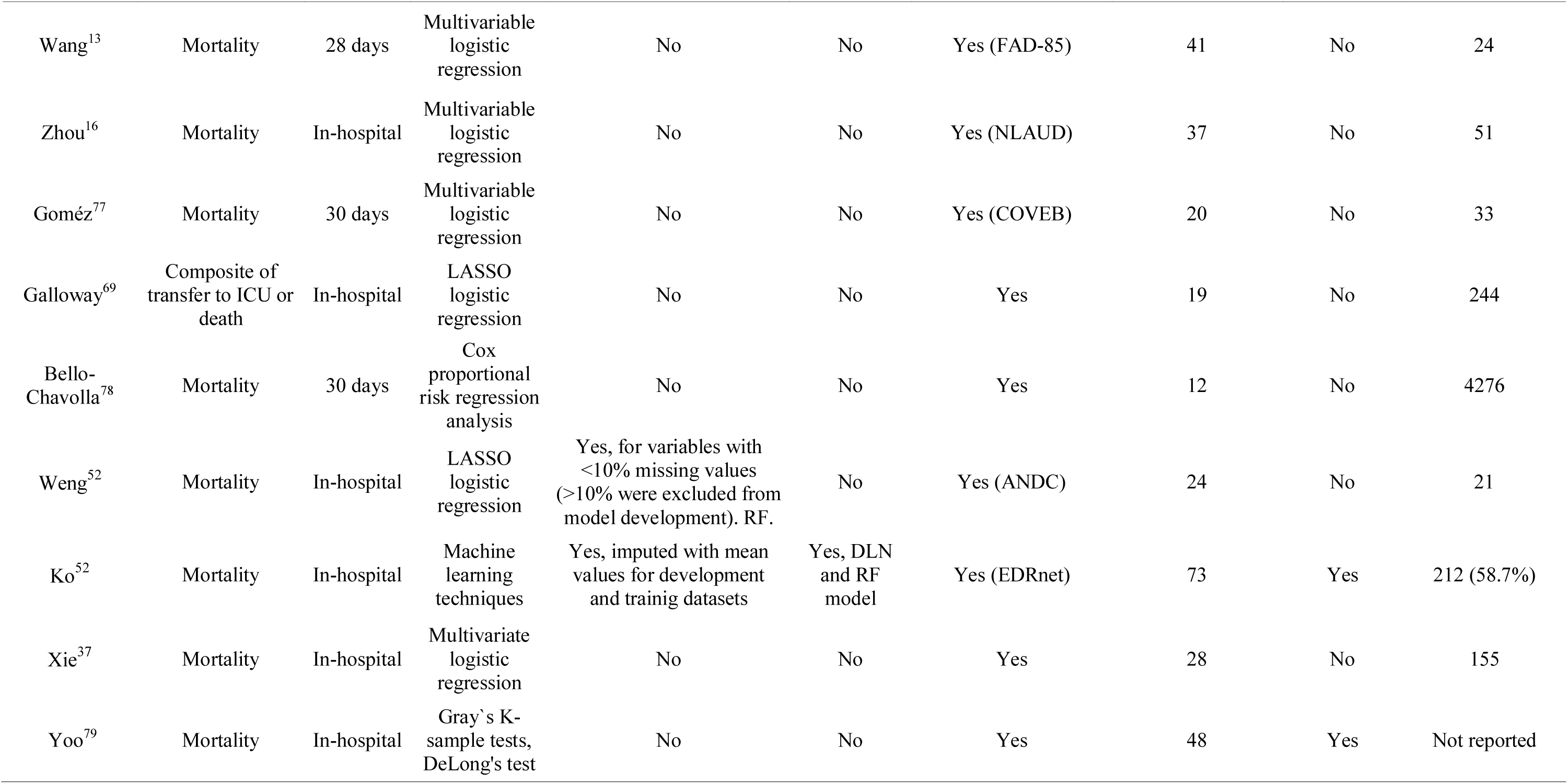
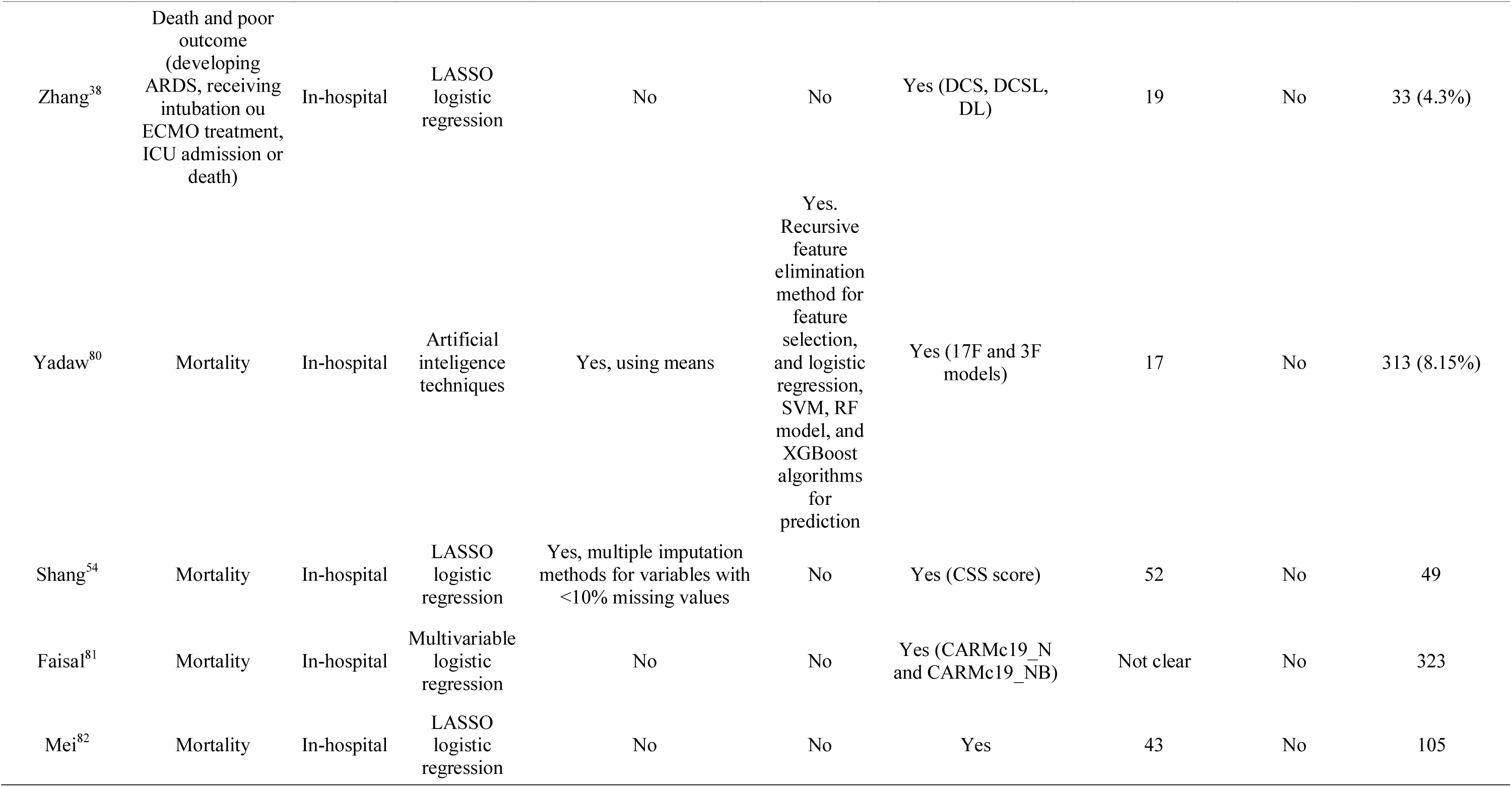
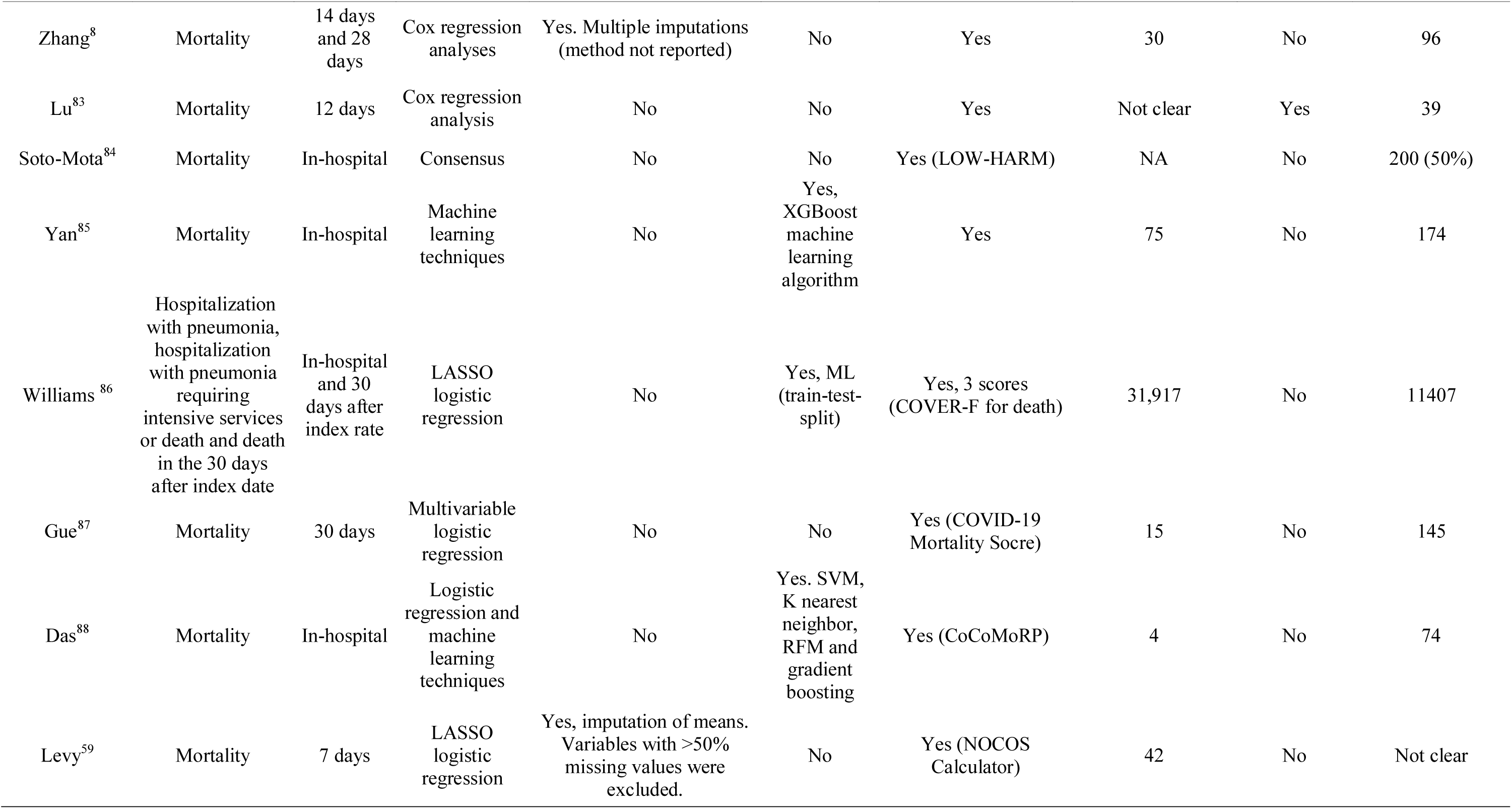
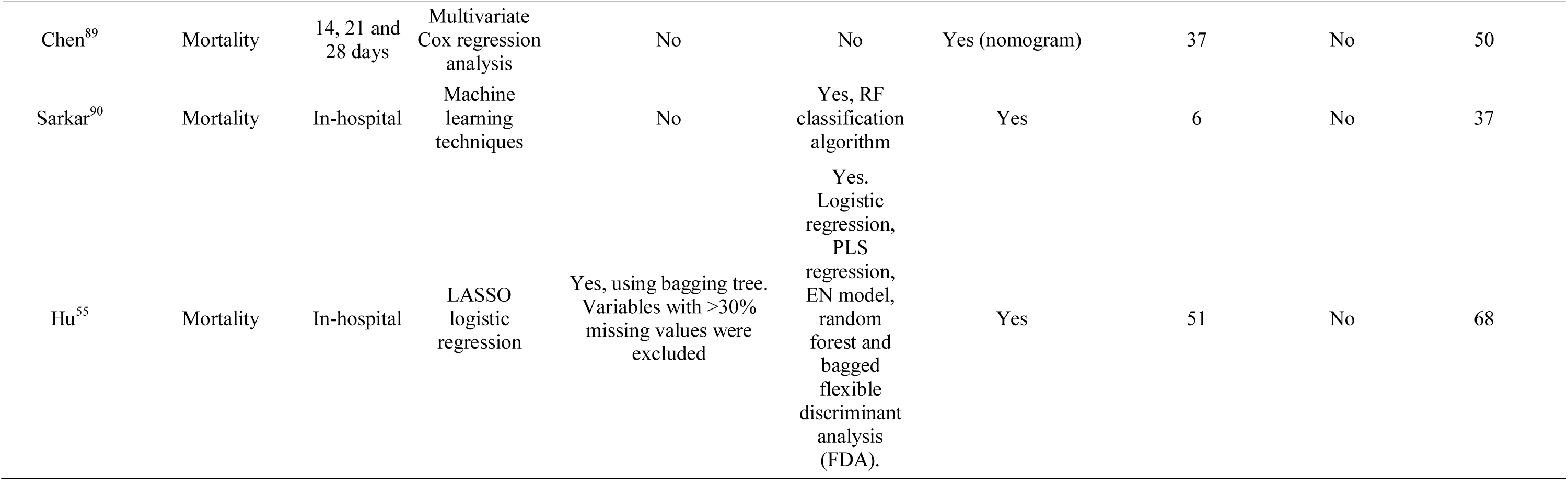
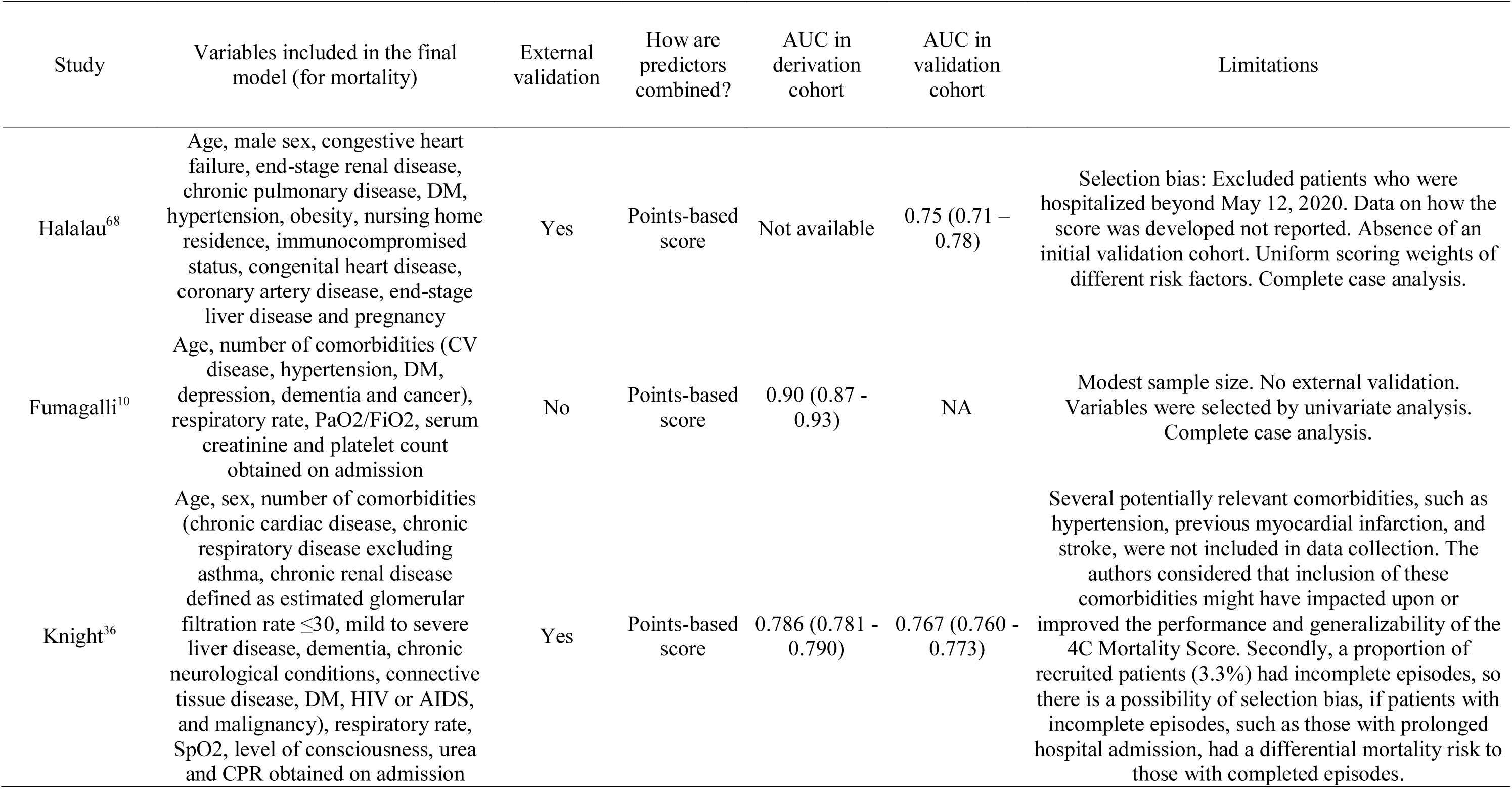
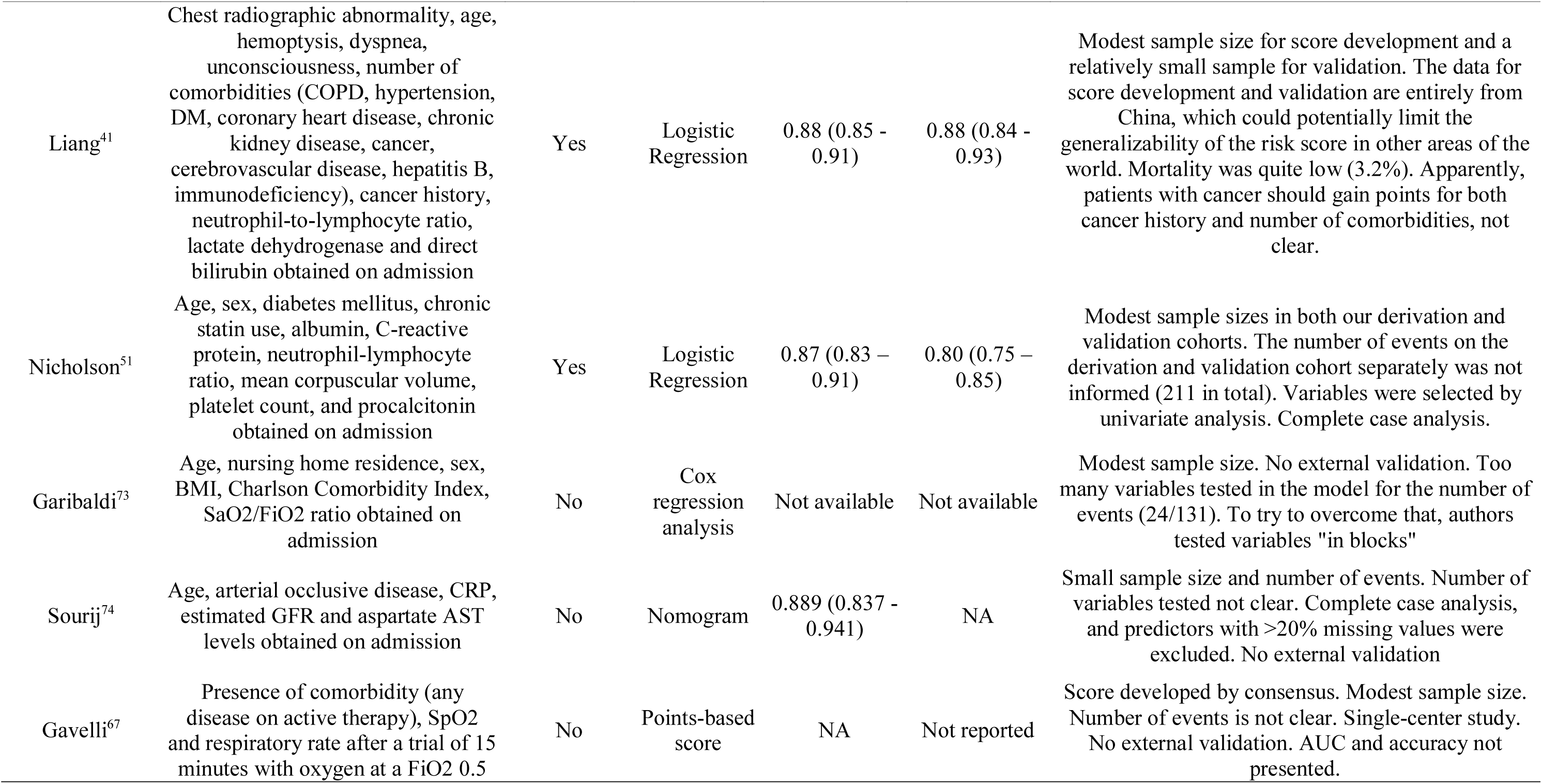
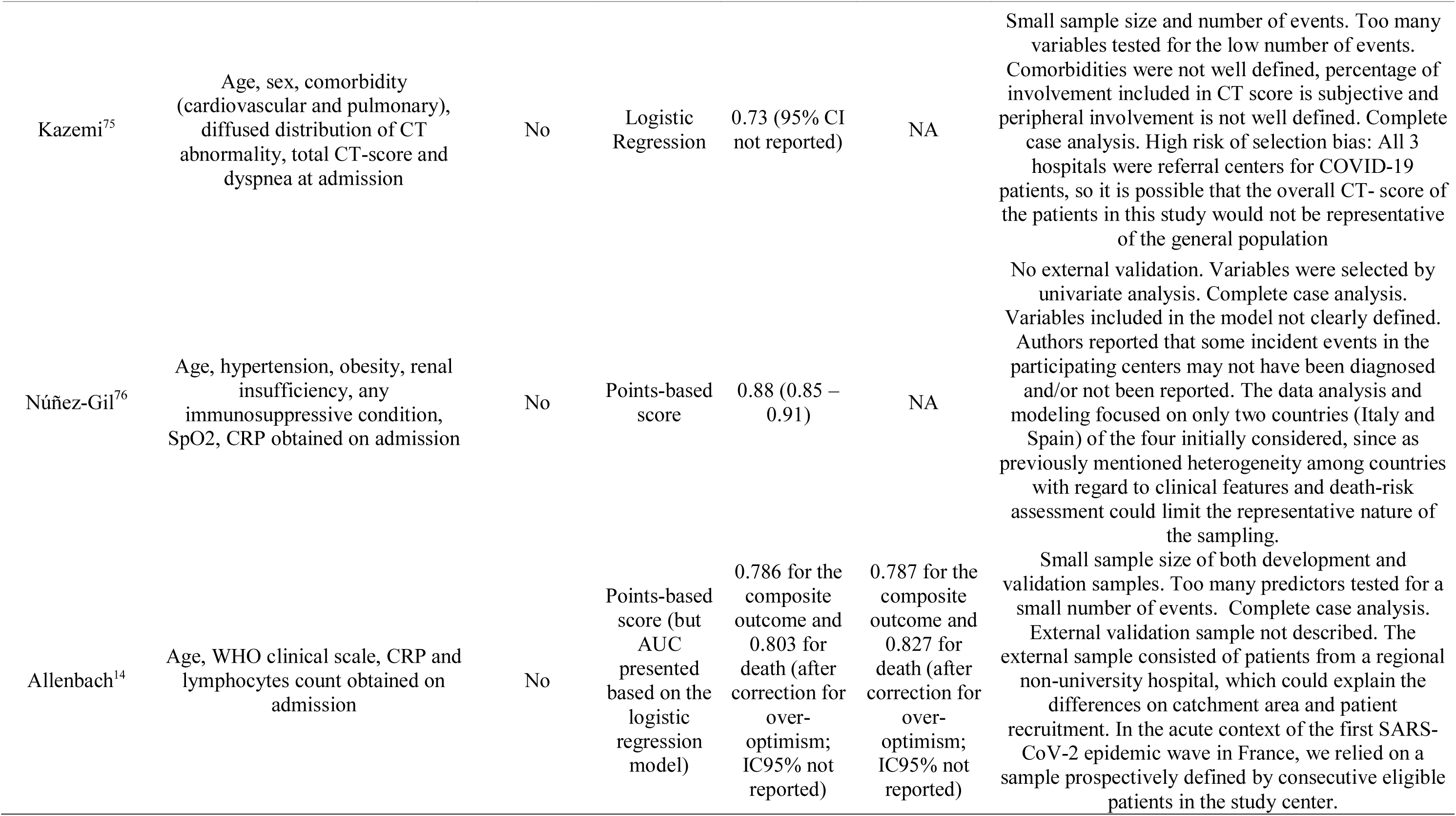
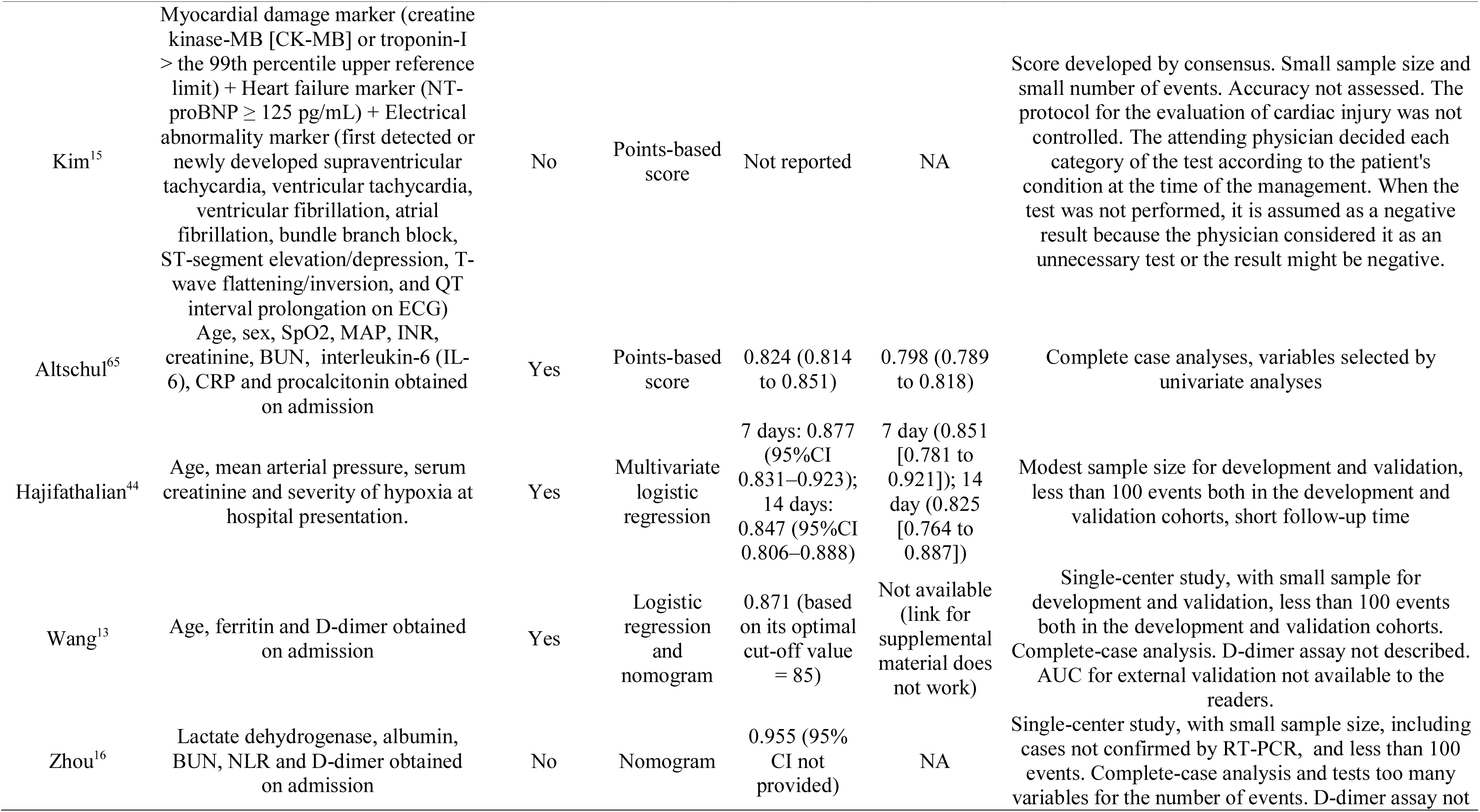
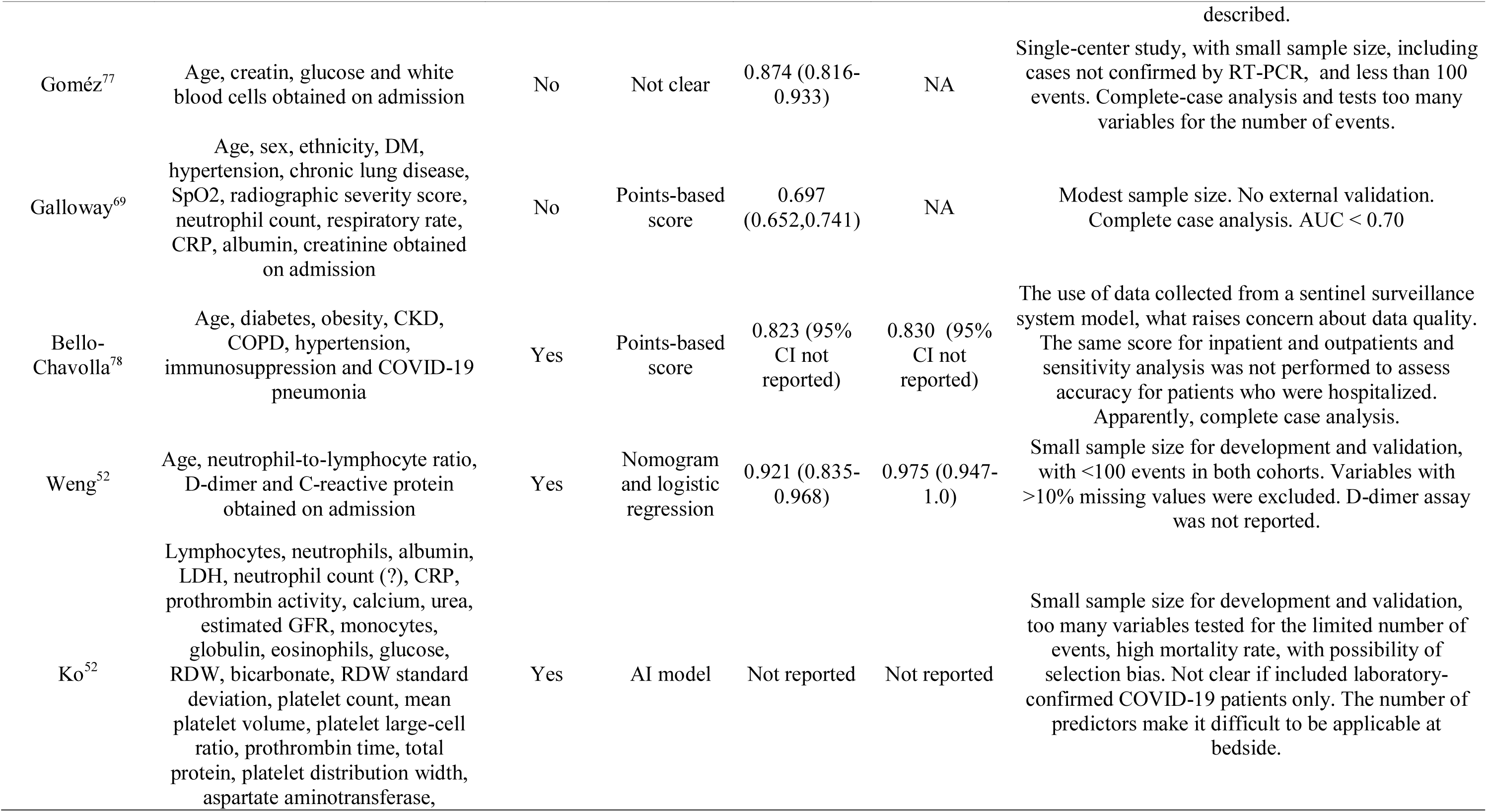
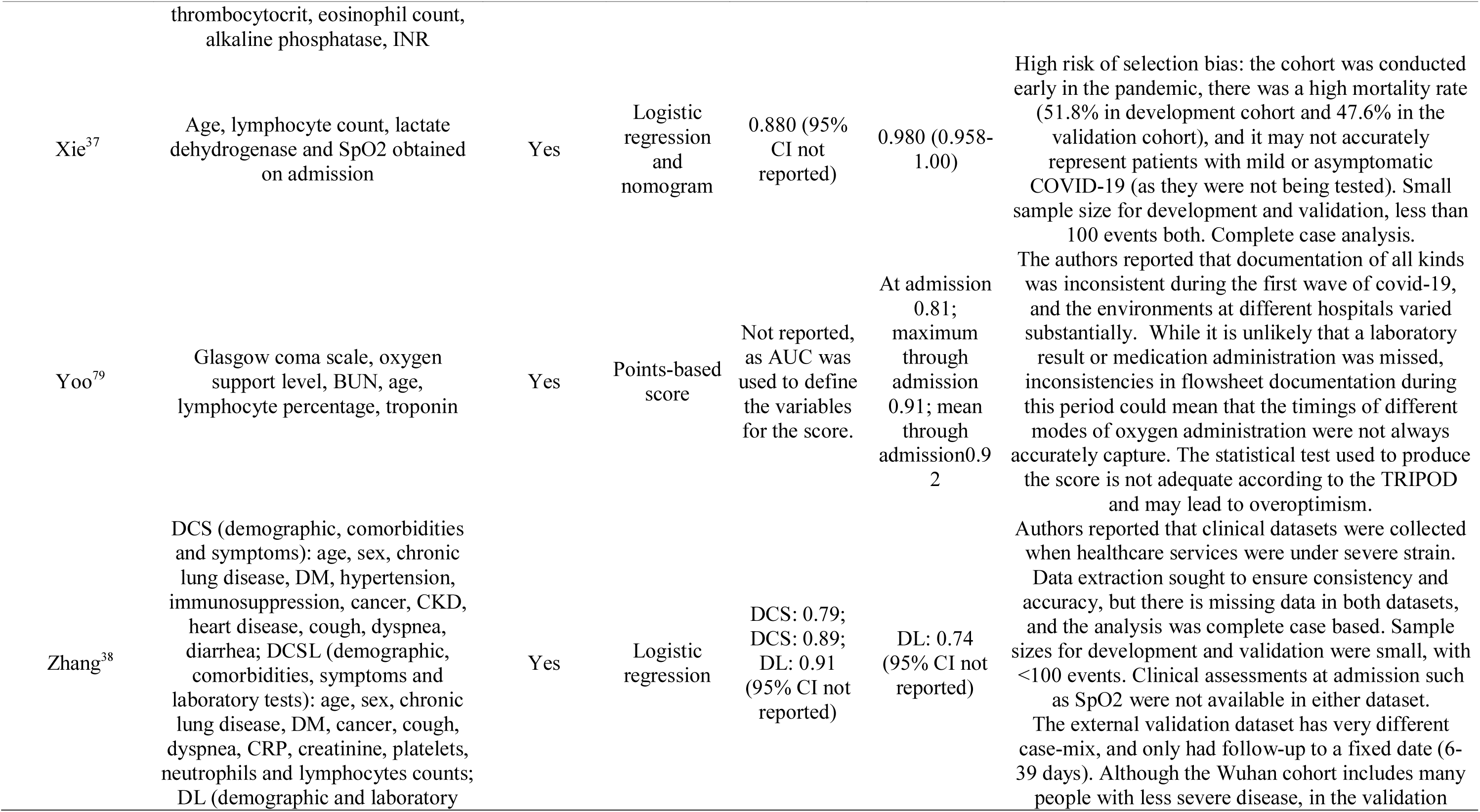
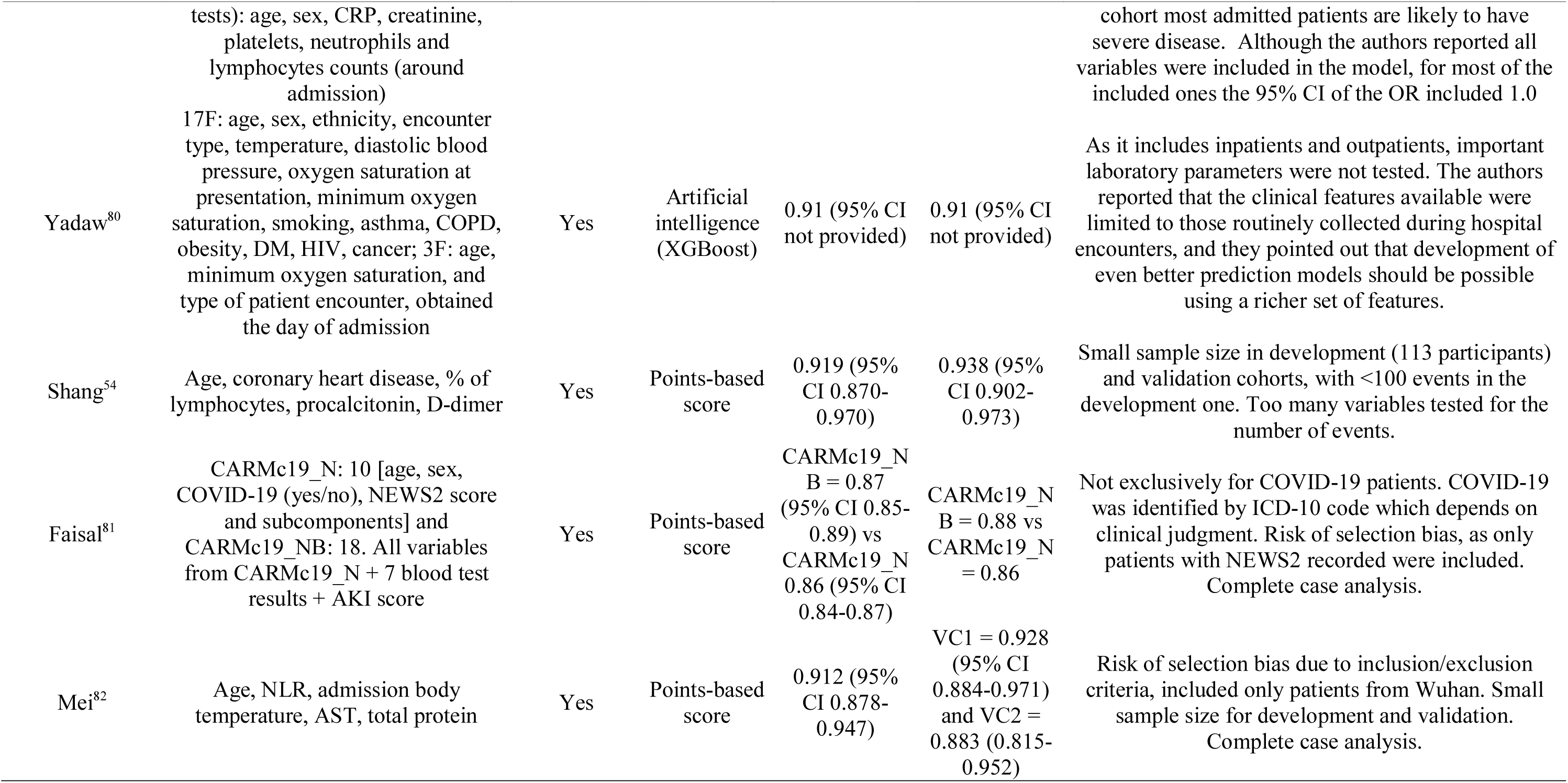
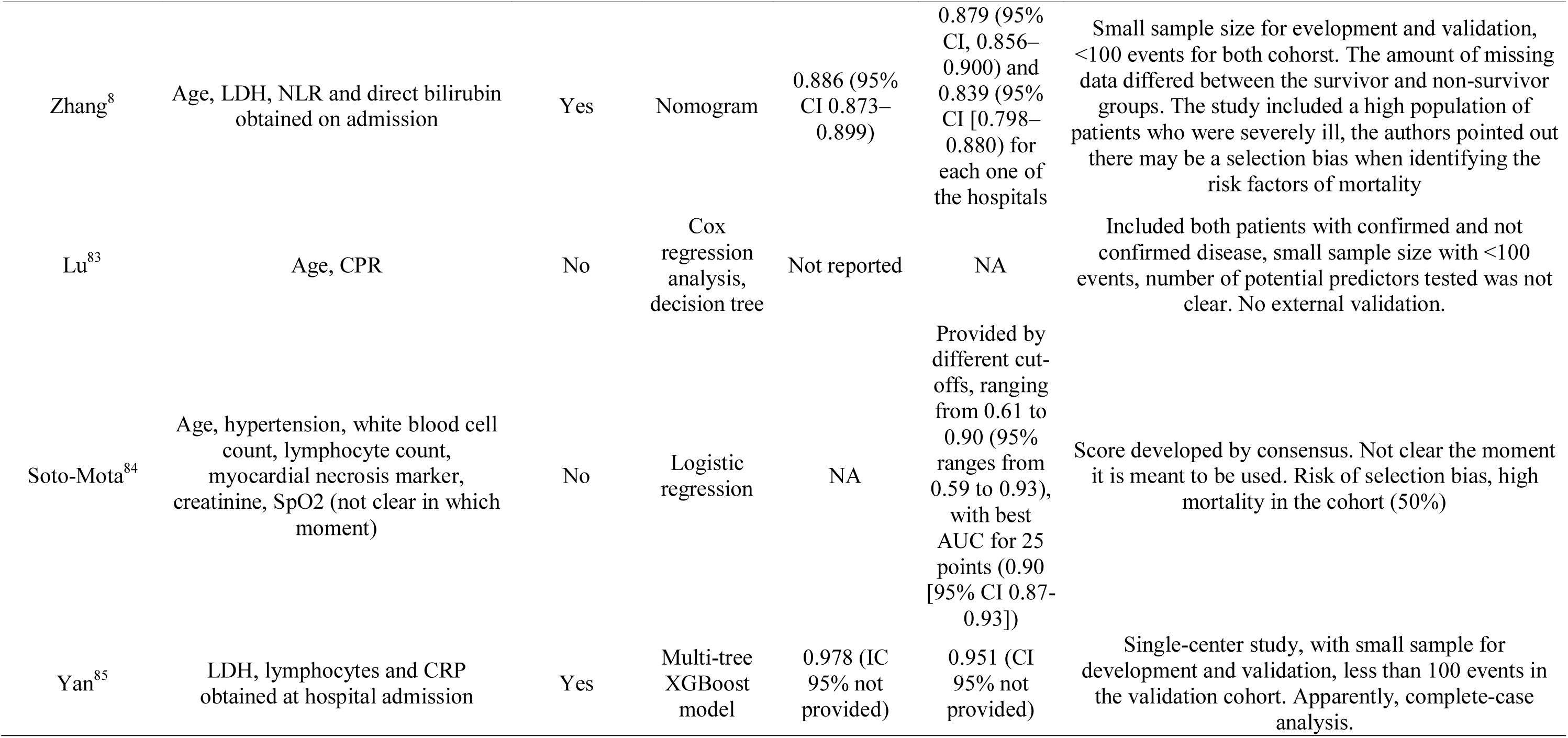
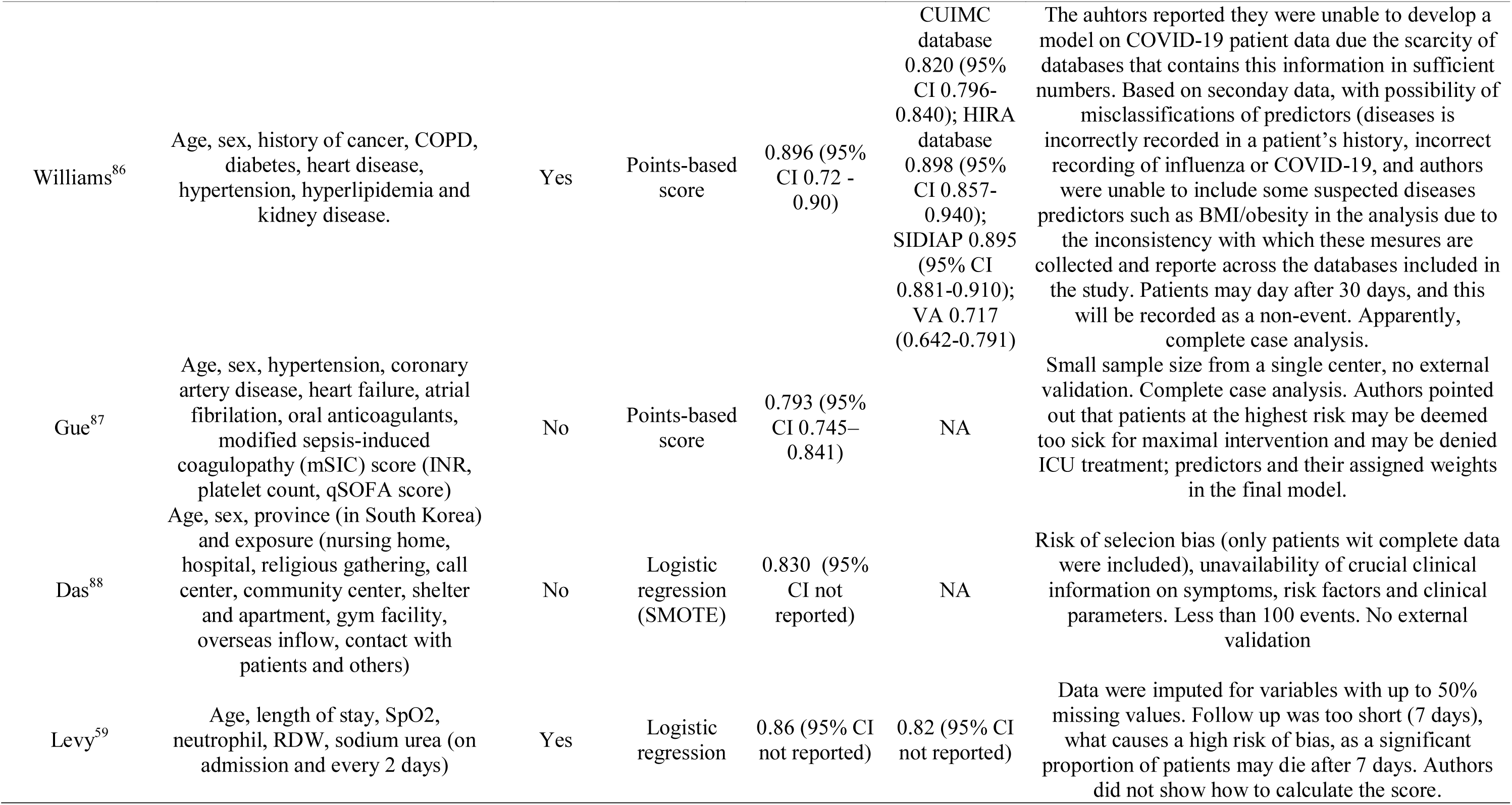
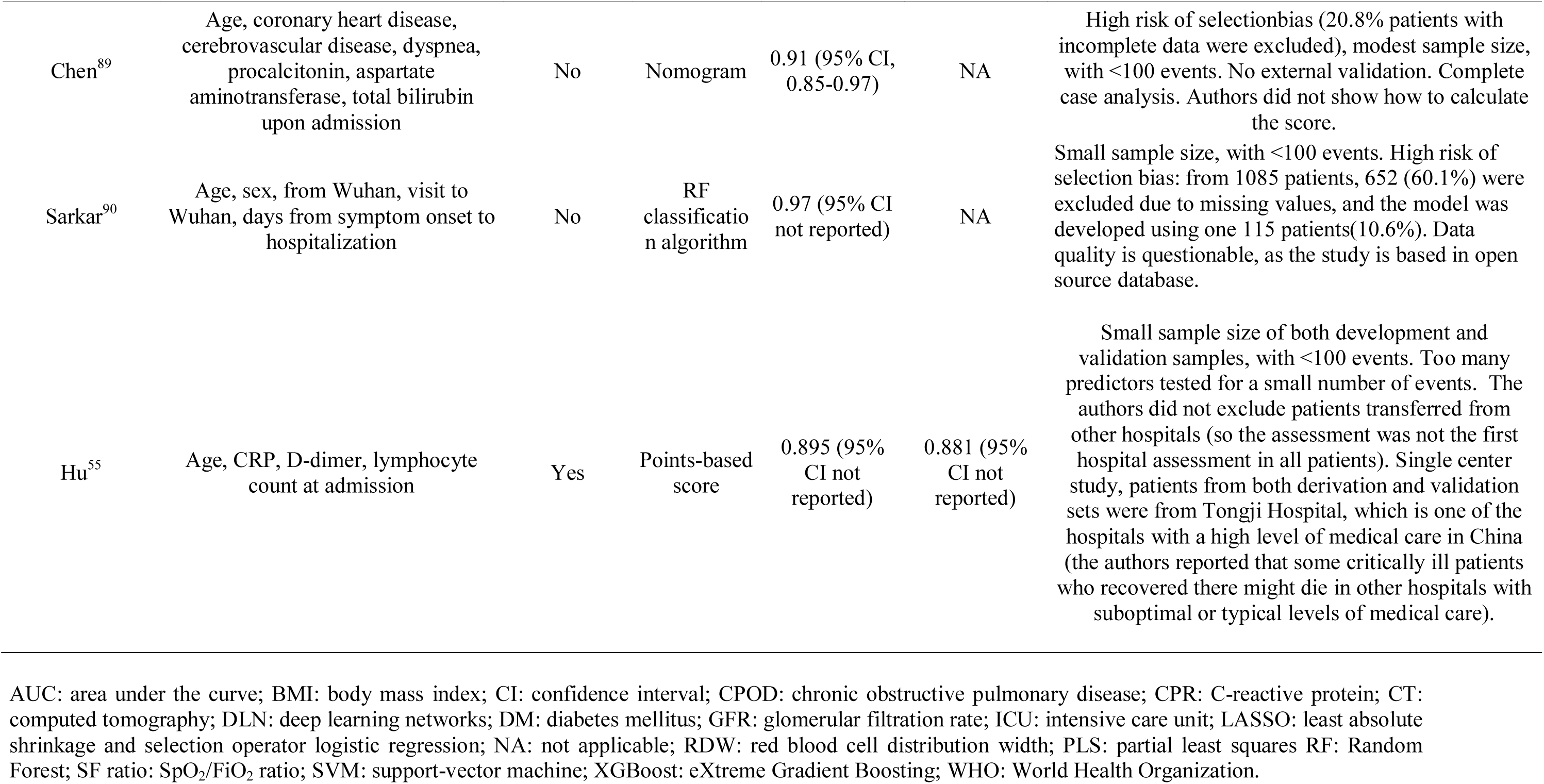
Main characteristics of the studies.

From the 27 (69.2%) developed scores to predict in-hospital mortality, in three studies the full information required for proper calculation was not available, in five studies the assay used for D-dimer or troponin was not described to allow proper comparison, in two studies the variables were not clearly defined (such as “kidney failure”, “elevated” CPR, and “cardiovascular and pulmonary comorbidity”), in two the variables were not applicable for other populations (such as province and coming from Wuhan), and in 12 one or more variables required were not in our study protocol.

### Comparison with other scores

Based on complete case validation cohort, the ABC_2_-SPH score achieved better discrimination (Table 6, Figure 5a) than other prediction scoring systems for COVID-19, pneumonia and sepsis (0.85; 95%CI: 0.82 – 0.88). Xie’s and Zhang’s score^8,37,38^ showed good discrimination, but the number of complete cases and deaths were relatively small. Considering clinical utility (Figure 5b), ABC_2_-SPH showed a better performance compared to the two most discriminating scores for in-hospital mortality that were tested in more than 700 patients (A-DROP and CURB-65^29^). COVID-AID-7 and COVID-AID-14 were not included, as they have assessed 7 and 14 day-mortality, respectively, and not in-hospital).

**Figure 5.**
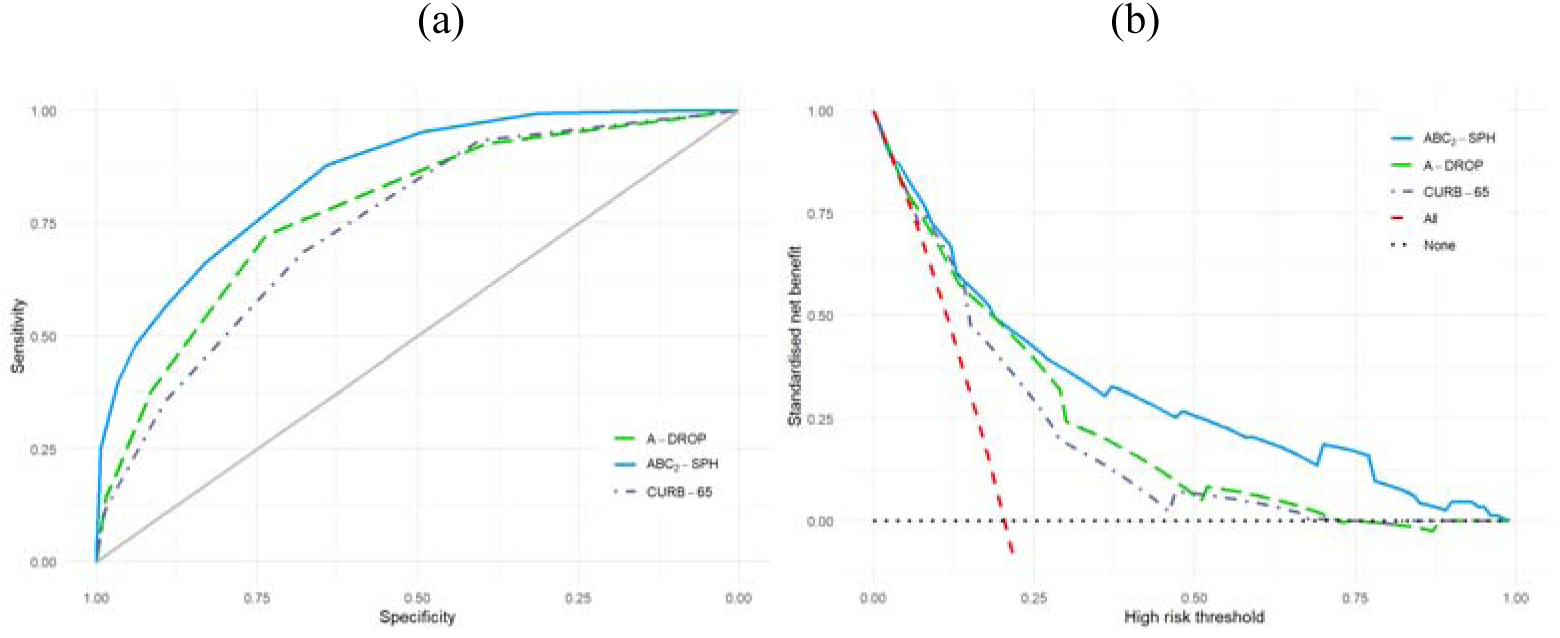
ROC curves (a) and decision curve for best performing scores.

**Table 6.**
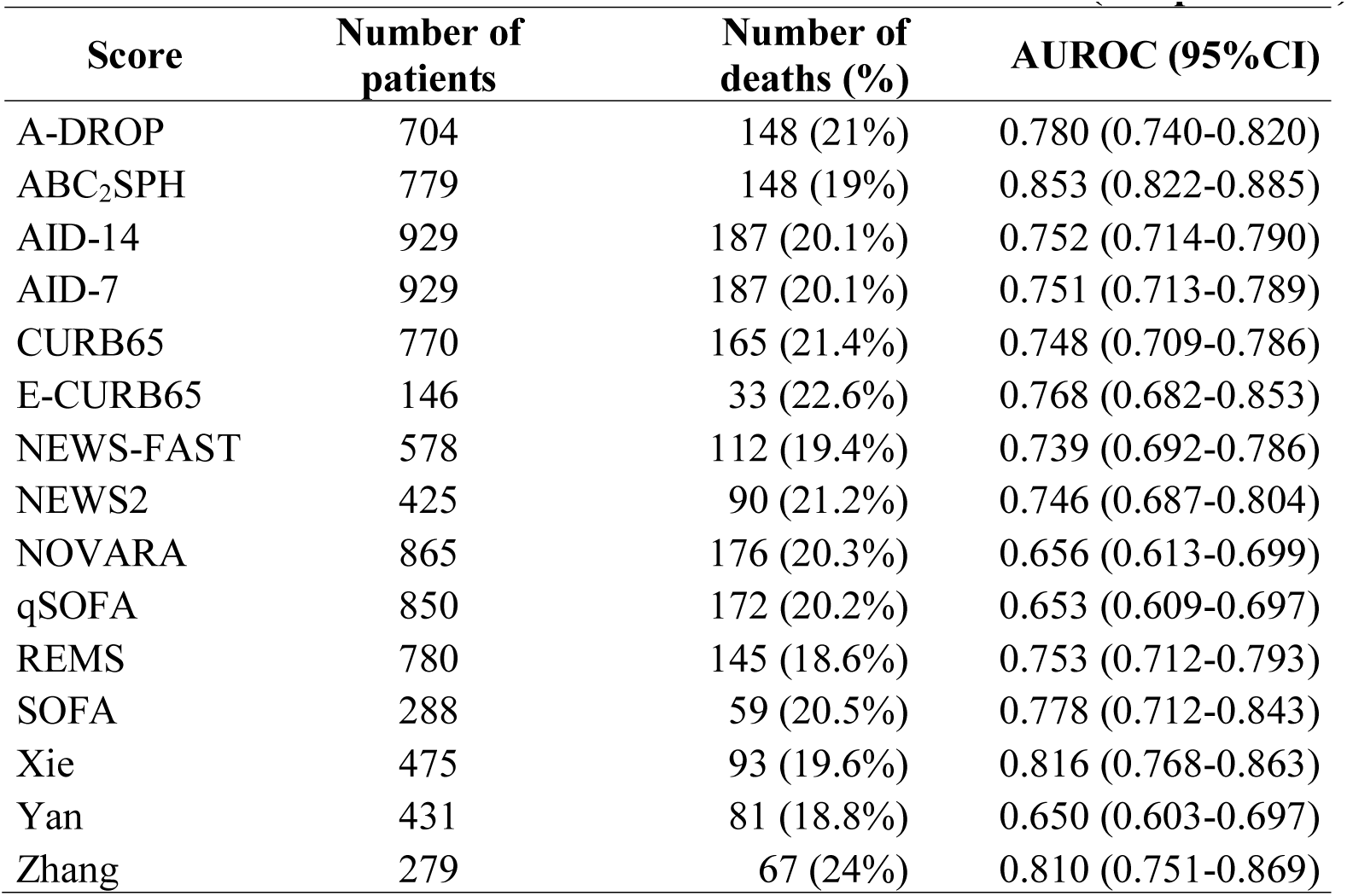
Discrimination of risk scores within validation cohort (complete case)

| **Score** | **Number of patients** | **Number of deaths (%)** | **AUROC (95%CI)** |
| --- | --- | --- | --- |
| A-DROP | 704 | 148 (21%) | 0.780 (0.740-0.820) |
| ABC<sub>2</sub>SPH | 779 | 148 (19%) | 0.853 (0.822-0.885) |
| AID-14 | 929 | 187 (20.1%) | 0.752 (0.714-0.790) |
| AID-7 | 929 | 187 (20.1%) | 0.751 (0.713-0.789) |
| CURB65 | 770 | 165 (21.4%) | 0.748 (0.709-0.786) |
| E-CURB65 | 146 | 33 (22.6%) | 0.768 (0.682-0.853) |
| NEWS-FAST | 578 | 112 (19.4%) | 0.739 (0.692-0.786) |
| NEWS2 | 425 | 90 (21.2%) | 0.746 (0.687-0.804) |
| NOVARA | 865 | 176 (20.3%) | 0.656 (0.613-0.699) |
| qSOFA | 850 | 172 (20.2%) | 0.653 (0.609-0.697) |
| REMS | 780 | 145 (18.6%) | 0.753 (0.712-0.793) |
| SOFA | 288 | 59 (20.5%) | 0.778 (0.712-0.843) |
| Xie | 475 | 93 (19.6%) | 0.816 (0.768-0.863) |
| Yan | 431 | 81 (18.8%) | 0.650 (0.603-0.697) |
| Zhang | 279 | 67 (24%) | 0.810 (0.751-0.869) |

## Discussion

### Main findings

ABC_2_-SPH score is simple, objective, easily available at hospital admission and easily calculated, employing seven well defined and routinely recordable variables: **a**ge, **b**lood urea nitrogen, number of **c**omorbidities, **C**-reactive protein, **S**pO2/FiO2 ratio, **p**latelet count, **h**eart rate. It has shown to be a reliable tool to estimate in-hospital mortality in COVID-19 patients, although true mortality risk is somewhat underestimated in very high risk patients. Model performance compared surpassed other existing scores.

The pandemic of COVID-19 disease has inflicted a heavy burden on the healthcare system of numerous countries. Little is known about how long the immune system will remain protective after vaccination or recovery from infection, and scientists have been predicting that SARS-CoV-2 “is here for the long haul”^39^. Therefore, it is of utmost importance to better identify those patients with higher risk of mortality, to inform early interventions and the need of more frequent repetitive assessments, to reduce the risk of death.

### Comparison with other studies

The majority of developed scores are limited by methodological bias in development cohorts.

Our prediction model was developed based on a large sample size of consecutive adult patients with confirmed COVID-19, from hospitals of different sizes, types and locations, to minimize the selection bias. Robust models require large sample sizes, which produce more reliable and accurate results^19^. It is estimated that 10-15 outcome events per predictor are required^40^. Among the models analyzed for comparison, only 30.8% used a sample with more than 1000 patients, 41.0% used a sample with less than 500 patients, and 41.0% were developed and validated in a sample with less than 100 events.

All studies have missing data^19^, this reality may be due to lack of standardization of the necessary exams at hospital admission, and differences in resources available in hospitals for carrying out tests. The approach of excluding the missing data and performing the analysis with the complete cases can lead to biased results, since the complete cases may not adequately represent the entire original study sample, generating a selection bias^19^. To avoid this type of bias, in our model multiple imputation with chained equations (MICE) was used to handle missing values on potential predictors, where the foul was assumed at random. Most of the models we analyzed for comparison (69.2%) did not perform or did not describe whether imputation methods were used for the missing data, therefore, there is a high risk of bias related to the treatment of missing data.

As the accuracy of a prediction model is always high whether the model is validated on the development cohort used to derive the model only, the assessment of accuracy in those studies may be overoptimistic. It is important that studies that develop prediction models use some validation method to quantify any optimism in the predictive performance of the model developed and adjust the model for overfitting^19^. In 43.6% of the analyzed studies, external validation of the developed model was not performed. External validation is highly recommended to assess the performance of a prediction model on other participant data that was not used for the development of the model^19^.

Previous studies have observed the variables included in the ABC_2_-SPH score as risk factors for severe COVID-19, what shows that our results are in line with the available evidence. Age and number of comorbidities were reported as independent risk factors for developing severe COVID and mortality in several publications^10,36,41^. The strong age gradient per decade after 60 years-old is in line with other series^10,14^. One of the main causes of death in COVID-19 patients is the unregulated immune response, with an uncontrolled production and secretion of cytokines. Aging is associated with a well-known decline in the adaptive and innate immunity, which plays a major role in the increased susceptibility of infections^42^. Age-related immune imbalance is also related to an increased severity in pro-inflammatory response and increased cytokine production, what is believed to increase patient vulnerability to the unregulated inflammatory response in COVID-19^43^. Other authors hypothesize that decreased lung elasticity, increased end-expiratory and abnormal alveolar integrity related to lung senescence, which may be associated to kidney senescence play a role in the predisposition for severe COVID-19 and mortality^10^.

It is important to highlight the evidence of a decreased antibody production following immunization in the elderly, as well as shortened duration of protective immunity^43^. This might be the case for COVID-19 as well. Therefore, even being a priority group for the vaccine, this age group will probably remain a major risk factor for mortality.

The number of comorbidities indicates the importance of pre-existing conditions to the severity of COVID-19. Even though comorbidities are age-dependent factors, the number of comorbidities remained as an independent risk factor in the final model.

As we aimed to use variables easily available at ED admission of any institution, we opted to evaluate the ratio of peripheral oxygen saturation over the inspired fraction of oxygen (SpO_2_/FiO_2,_ SF ratio), instead of the ratio of arterial oxygen partial pressure over the fraction of inspired oxygen, like the COVID-AID score^44^. Arterial blood gas puncture and analysis is an invasive and complex procedure, which may be time consuming for the team. Additionally, a recent publication highlighted that despite widespread familiarity with use of PaO_2_/FiO_2_ ratio using blood gas analysis, clinical recognition of acute respiratory distress syndrome remains poor^45^. The authors assessed 28,758 mechanical SF ratio and PaO_2_/FiO_2_ ratio in mechanical ventilated patients, and observed that PaO2/FiO_2_ ratios were substantially less available or even unavailable in a significant proportion of ventilated patients^45^. SF ratio was already validated as a substitute for the PaO_2_/FiO_2_ ratio in assessing the oxygenation criterion of patients with acute lung injury and acute respiratory distress syndrome^46^.

COVID-19 associated hyperinflammation and coagulopathy are correlated to with a wide deviation in various inflammatory markers and hemostasis parameters, including C-reactive protein, thrombocytopenia, D-dimer and prothrombin time, and thus these are potential prognostic markers of increased mortality in COVID-19^47,48^.

Consistent with prior studies, we also observed utility of CRP thrombocytopenia.

C-reactive protein in an acute phase reactor with established prognostic prediction role in intensive care septic and non-septic patients^49,50^, and it has been included in different scores an independent predictor for mortality.^51,52^

The prognostic value of thrombocytopenia in patients with COVID-19 has shown in a recent meta-analysis^53^, and it was also included in other scores^10,51^. The exact explanation is still unknown, and it is probably multifactorial, related to direct infection of bone marrow cells by the virus, resulting in abnormal hematopoiesis; platelet destruction by the immune system; endothelial damage triggering platelet activation, aggregation and microthrombi in the lung; and abnormal platelet defragmentation in the lungs^53^.

A recent meta-analysis of 16 studies, which included 2783 surviving and 697 non-surviving cases, has shown significantly higher levels of D-dimer on admission in patients who died compared to the ones who were discharged^47^. This exam was included as a predictor in different scores^13,16,52,54,55^. Although D-dimer was collected in our study, D-dimer assays varied widely among different hospitals. Ideally, the value has to be determined with the same methodology, preferably from the same manufacturer, and this information was not available in any of the studies.

A recent publication highlighted confusion and potential for misinformation in reporting D-dimer data in COVID-19^56^. The authors emphasized that the considerable variation in reporting units for D-dimer is potentially under-recognized in various studies, with at least 28 potential theoretical combinations of measuring units for D-dimer, either D-dimer units (DDU) or fibrinogen equivalent units (FEU), which are approximately 2× those of DDU. There is also possibility for misreporting of D-dimer data based on poor or incomplete reporting. The authors provided examples of serious errors in the reported values and/or units as reported in the literature related to COVID-19, even in high impact journals.

Most studies have not reported how they dealt with cases who were transferred between hospitals. Although SOFA scores tend to be low at hospital admission, Zhou et al^57^ observed that age, SOFA score and D-dimer at admission were independent risk factors for mortality. However, they opted to include those patients, even patients with late stage COVID-19, using admission data from the second hospital only. It is quite likely there was a higher chance those patients were already with critical disease^57^. As the score is intended to be used at hospital admission, we opted to exclude patients who were transferred between hospitals and admission data from the first hospital was not available.

Blood urea nitrogen elevation was a strong predictor for mortality, what is in line with other scores^36,58,59^. Kidney disease has been widely described as a risk factor for in-hospital mortality. Although autopsy studies did not find conclusive evidence of SARS-CoV2 infection in the kidney, some authors hypothesize that the damage may be mediated by direct cytopathic effects of SARS-Cov2 on the kidney tissue, immune-mediated damage due to virus-induced immune complexes, as well as the effects of the inflammatory response, hypoxia and shock^60–62^.

### Strengths and limitations

A major strength of this rapid scoring system is its simplicity, the use of objective parameters, what helps to reduce inter-user variability, easily available at the emergency department presentation, even in under-resourced settings. A major strength of this study is that it followed strict methodological criteria, recommended by TRIPOD checklist and PROBAST^20^, and was based on robust sample of patients with laboratory confirmed SARS-CoV-2 infection, from a collaboration among researchers from 36 public, private and mixed hospitals of different sizes in four Brazilian states, to ensure diversity of the population studied and representativeness of the intended target population. The majority of published scores were developed in China or the US (56.4%) and Europe (25.6%), this is the first study in the Latin American population. Data were obtained by detailed medical chart reviews, and we were able to collect comprehensive data from a large number of patients and follow 98.5% of the patients from admission to discharge or death. Decisions about which predictors to retain in the final model did not rely on potentially biased univariable selection of predictors. They were based on clinical reasoning, previous evidence from other cohorts and systematic reviews on prognostic factors for COVID-19 patients and availability of predictor measurement at hospital admission^19^.

In a huge country such as Brazil, the development of a score that truly corresponds to the reality of our population’s characteristic was only possible by the collaborative work among several hospitals from all the regions of the country. The COVID-19 cause and requirement for agile answers from the scientific community motivated the fast and precise teamwork and allowed the achievement of the creation of a tool to support the daily work in the frontline to combat the pandemic. We believe that the learning regarding the development of qualified and useful research engaging several centers could allow us to generate more accurate and faster results to subside health policies in the future.

Patients who were transferred to other hospitals and thus were lost to follow-up do not characterize selection bias, as they similar characteristics of the development and validation cohorts, and a risk similar to those cohorts: of the 77 patients, 53 presented complete data and had their scores calculated - low risk 30.2%, intermediate 35.8%, high 22.6%, very high 11.3%.

With regards to study limitations, it was a retrospective analysis subject to the drawbacks of patient records review. Obesity was not directly measured by body mass index, but rather clinical defined, gathered from medical records, which may have led to underreporting. Due to the pragmatic study design, laboratory tests were performed at the discretion of the treating physician, and we did not have a full dataset on all laboratory parameters of interest available. Some laboratory parameters, which proved to be of prognostic relevance in other studies, were not available for at least 2/3 of patients in our sample. Therefore, we cannot rule out that variables with a higher proportion of missing data would have had a significant impact on mortality prediction. Additionally, we were unable to assess the predictive ability of some scores, as some required variables were not available.

Another bias from the Spanish validation cohort is the fact that the majority of those patients came from the beginning of the pandemic, and management of the patients improved during subsequent waves. Data include 28-day mortality, which may differ from the Brazilian data, although the score was able to show very good discrimination.

### Implications for clinicians and policymakers

ABC_2_-SPH score may be very useful in a real-world setting, to provide healthcare practitioners the decision support that is needed to help them better identify and prioritize the care of patients who have the higher risk of death. Its development and validation followed strict methodological criteria, and the score fulfils the majority of the characteristics of an ideal score^63^. It can be used in all emergency departments, regardless of the level of resource settings. The results represent the experience of 36 hospitals in 17 cities in Brazil, and one hospital in Spain, and they are highly relevant to the current pandemic. It can be easily calculated at bedside or could be easily integrated to the electronic medical records for an automatic computation. It may help clinicians to identify high-risk patients from the triage phase, as well as to identify those most appropriate to be enrolled into therapeutic trials, may make possible to expand inclusion criteria through the early identification of patients who may benefit from therapy^64^. It might also be useful to help guiding recommendations for early palliative consultation^65,66^.

Different from what has been mistakenly suggested^36,67–69^, the results from this study do not suggest that patients from low-risk group may be discharged for home treatment. No score so far has specifically tested this hypothesis. A recent editorial has highlighted the importance of taking into account the “treatment paradox”: patients identified to be at the low risk group were at low risk due to the interventions received in hospital^70^. It must not be interpreted as the risk to a patient if no actions are taken. Sperrin & McMillan counterfactual prediction modelling as a potential solution to minimize bias from treatment paradox^70^. More importantly, due to the treatment paradox, scoring systems developed and validated in in-hospital settings cannot be used in outpatient settings without further validation, as it has been mistakenly suggested^71^.

### Unanswered questions and future research

We believe that ABC_2_-SPH score may hold potential generalizability for other countries. However, prediction models are population specific and may produce different results in different populations^72^. Considering that thresholds for admission may vary, hospitalized COVID-19 population may be different, the outcome events are different and patient management may be different, further validation (and re-calibration) in different health care settings is recommended. In particular, we learned that our model might underestimate mortality in high-risk individuals.

As we opted to develop the score focusing on information available at admission, as this would make it more useful for clinicians, other important factors during hospitalization that may impact prognosis were not included. Further analysis involving these factors are required.

ABC_2_-SPH score may help clinicians to make a prompt and reasonable decision to optimize patient management and potentially reduce mortality. However, further prospective studies are needed to investigate whether the use of the score in the emergency department indeed trigger actions that result in reduced complications and hospital mortality. Additionally, due to the rapidly changing nature of the COVID-19 and the disease management, model performance should be monitored closely over time and space^70^.

Future studies may also investigate risk factors for mortality among patients who develop COVID-19 symptoms during hospital admission due to other conditions.

### Conclusion

In conclusion, we developed and validated the ABC_2_-SPH rapid scoring system and a web-based risk calculator. This score, based on age, number of comorbidities, blood urea nitrogen, C-reactive protein, platelet count, peripheral oxygen saturation and oxygen support at admission is an inexpensive tool, showed to objectively and accurately predict in-hospital mortality in COVID-19 patients. It may be used at bedside for earlier identification of in-hospital mortality risk and, thus, inform clinical decisions and the assignment to the appropriate level of care and treatment for COVID-19 patients.

## Data Availability

Data are available upon reasonable request.

## Acknowlegments

We would like to thank the hospitals which are part of this collaboration, for supporting this project: Hospital Bruno Born; Hospital Cristo Redentor; Hospital das Clínicas da Faculdade de Medicina de Botucatu; Hospital das Clínicas da UFMG; Hospital das Clínicas da Universidade Federal de Pernambuco; Hospital de Clínicas de Porto Alegre; Hospital de Santo Antônio; Hospital Eduardo de Menezes; Hospital João XXIII; Hospital Julia Kubitschek; Hospital Mãe de Deus; Hospital Márcio Cunha; Hospital Mater Dei Betim-Contagem; Hospital Mater Dei Contorno; Hospital Mater Dei Santo Agostinho; Hospital Metropolitano Dr. Célio de Castro; Hospital Metropolitano Odilon Behrens; Hospital Moinhos de Vento; Hospital Nossa Senhora da Conceição; Hospital Regional Antônio Dias; Hospital Regional de Barbacena Dr. José Américo; Hospital Regional do Oeste; Hospital Risoleta Tolentino Neves; Hospital Santa Cruz; Hospital Santa Rosália; Hospital São João de Deus; Hospital São Lucas da PUCRS; Hospital Semper; Hospital SOS Cárdio; Hospital Tacchini; Hospital Unimed-BH; Hospital Universitário Canoas; Hospital Universitário Ciências Médicas; Hospital Universitário de Santa Maria.

We also thank all the clinical staff at those hospitals, who cared for the patients, and all undergraduate students who helped with data collection.

## Data availability statement

Data are available upon reasonable request.

## Transparency declaration

The lead authors (MSM and MCP) affirm that the manuscript is an honest, accurate, and transparent account of the study being reported; that no important aspects of the study have been omitted; and that any discrepancies from the study as originally planned (and, if relevant, registered) have been explained.

---

**Table S1.** Demographic and clinical characteristics for patients admitted to hospital with COVID-19 and were transferred to other hospitals (n=77)

| **Characteristic** | **Frequency (%) or median (IQR)** | **Non missing cases (%)** |
| --- | --- | --- |
| Age (years) | 55.0 (51.0, 70.0) | 77 (100%) |
| Sex at birth |  | 77 (100%) |
| Male | 48 (62.3%) |  |
| **Comorbidities** |  |  |
| Hypertension | 41 (53.2%) | 77 (100%) |
| Coronary artery disease | 4 (5.2%) | 77 (100%) |
| Heart failure | 5 (6.5%) | 77 (100%) |
| Atrial fibrillation or flutter | 2 (2.6%) | 77 (100%) |
| Stroke | 3 (3.9%) | 77 (100%) |
| COPD | 4 (5.2%) | 77 (100%) |
| Diabetes mellitus | 22 (28.6%) | 77 (100%) |
| Obesity (BMI>30kg/m<sup>2</sup>) | 8 (10.4%) | 77 (100%) |
| Cirrhosis | 2 (2.6%) | 77 (100%) |
| Cancer | 5 (6.5%) | 77 (100%) |
| Number of comorbidities |  | 77 (100%) |
| 0 | 23 (29.9%) |  |
| 1 | 24 (31.2%) |  |
| 2 | 20 (26.0%) |  |
| 3 | 8 (10.4%) |  |
| 4 | 2 (2.6%) |  |
| **Clinical assessment at admission** |  |  |
| SF ratio | 433.3 (350.0, 447.6) | 75 (97.4%) |
| Respiratory rate (irpm) | 22.0 (18.0, 24.0) | 61 (79.2%) |
| Heart rate (bpm) | 89.0 (78.2, 99.8) | 70 (90.9%) |
| Glasgow coma score | 15.0 (15.0, 15.0) | 75 (97.4%) |
| Systolic blood pressure (mmHg) |  | 70 (90.9%) |
| < 90 | 2 (2.9%) |  |
| ≥ 90 | 68 (97.1%) |  |
| Diastolic blood pressure |  | 70 (90.9%) |
| ≤ 60 | 12 (17.1%) |  |
| > 60 | 58 (82.9%) |  |
| Inotrope need at admission | 0 (0%) |  |
| Laboratory |  |  |
| Hemoglobin (g/L) | 13.6 (12.2, 14.9) | 71 (92.2%) |
| Platelet count (10<sup>9</sup>/L) | 196.0 (144.0, 250.0) | 71 (92.2%) |
| Neutrophils-to-lymphocytes ratio | 5.7 (4.0, 8.4) | 62 (80.6%) |
| Lactate (mmol/L) | 1.3 (1.1, 1.9) | 45 (58.4%) |
| C-reactive protein (mg/L) | 87.5 (61.2, 134.5) | 62 (80.6%) |
| BUN (mg/dL) | 41.0 (19.1, 28.5) | 69 (89.6%) |
| Creatinine (mg/dL) | 1.1 (0.8, 1.4) | 73 (94.8%) |
| Sodium (mmol/L) | 138.0 (135.0, 141.0) | 65 (84.4%) |
| Bicarbonate (mEq/L) | 21.9 (20.0, 23.2) | 59 (76.6%) |
| pH | 7.4 (7.4, 7.5) | 60 (77.9%) |
| pO<sub>2</sub> (mmHg) | 78.0 (62.1, 99.7) | 59 (76.6%) |
| pCO<sub>2</sub> (mmHg) | 32.0 (27.9, 35.5) | 59 (76.6%) |

BMI: body mass index; BUN: blood urea nitrogen; COPD: chronic obstructive pulmonar disease; SF ratio: SpO<sub>2</sub>/FiO<sub>2</sub> ratio.

**Table S2.**
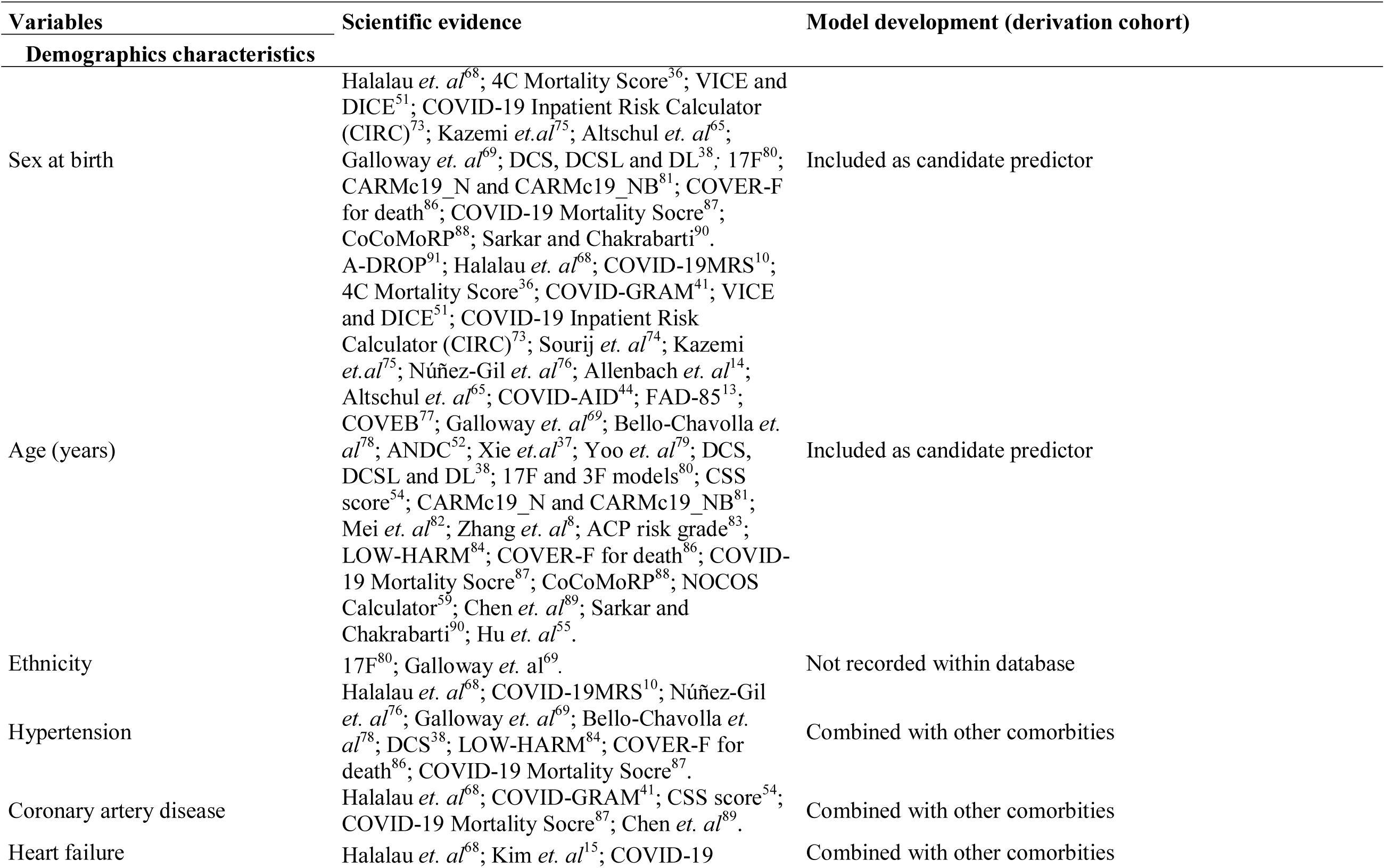
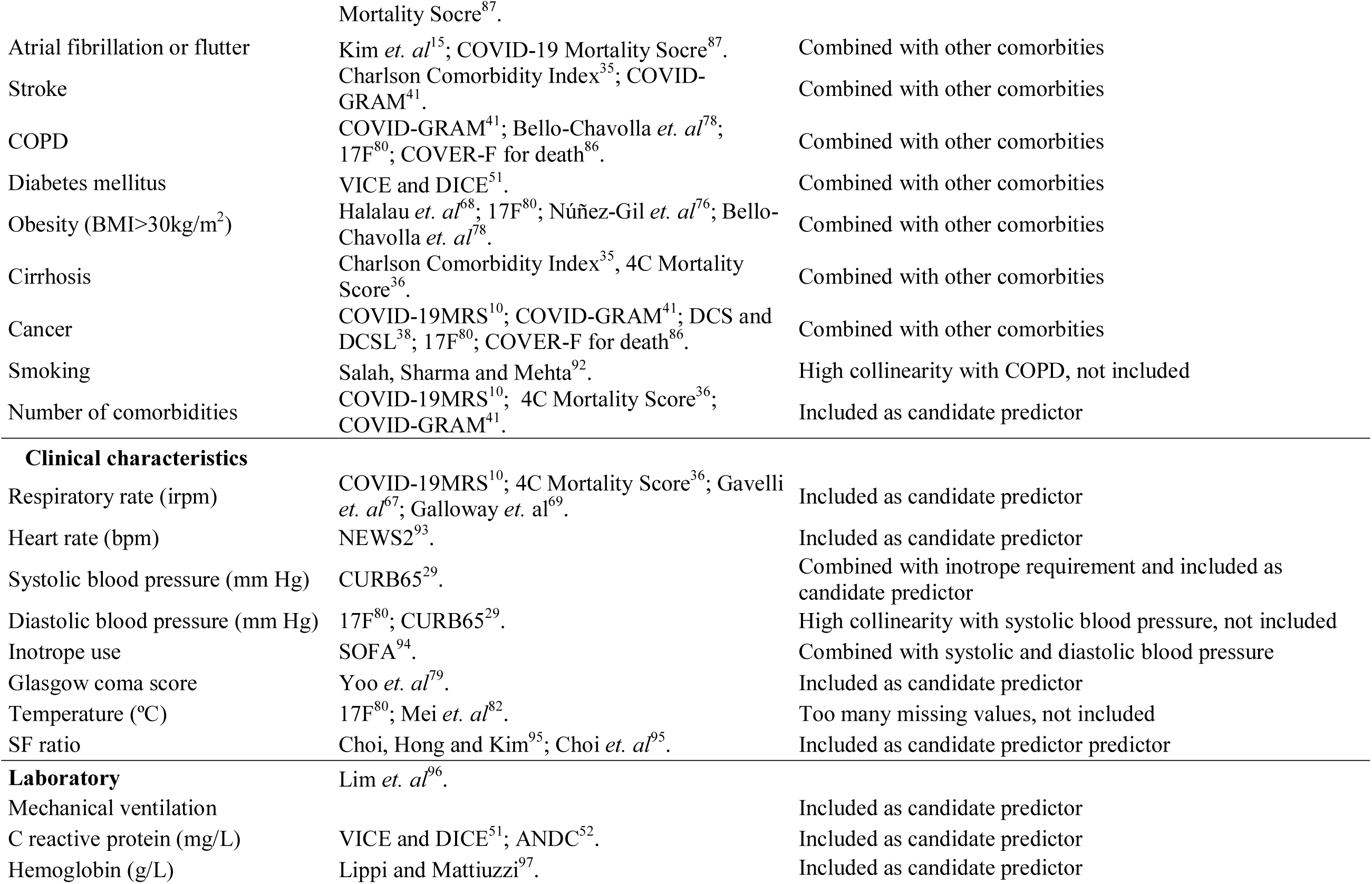
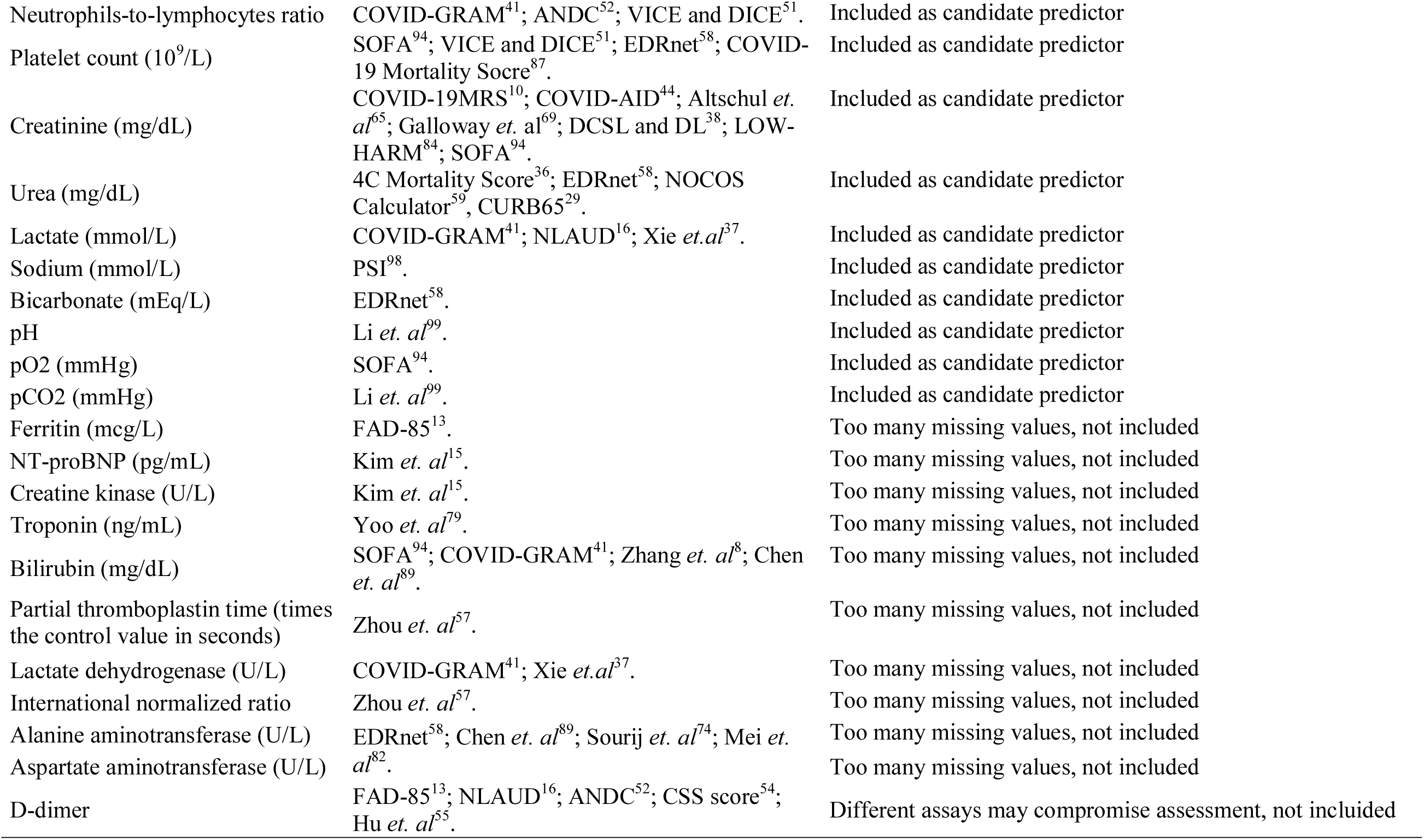
Evaluating potential predictors for the model development.

**Table S3.** Variable selection based on Generalized Additive Model.

| Variable | Deviance explained (%) | R-sq.(adj) | UBRE | D1-statistics (p-value) | D2-statistics (p-value) |
| --- | --- | --- | --- | --- | --- |
| All variables included | 0.354 | 0.361 | -0.324 |  |  |
| Sex at birth | 0.354 | 0.361 | -0.325 | 0.773 | 0.785 |
| Age (years) | 0.314 | 0.320 | -0.284 | 0.000** | 0.000** |
| Number of comorbidities | 0.353 | 0.361 | -0.323 | 0.011** | 0.011** |
| Respiratory rate (irpm) | 0.351 | 0.358 | -0.321 | 0.246 | 0.131 |
| Heart rate (bpm) | 0.350 | 0.357 | -0.320 | 0.047** | 0.122 |
| Systolic blood pressure (mm Hg) | 0.353 | 0.361 | -0.324 | 0.217 | 0.244 |
| Glasgow coma score | 0.353 | 0.360 | -0.324 | 0.995 | 1.000 |
| SF ratio | 0.333 | 0.339 | -0.303 | 0.000** | 0.000** |
| C-reactive protein (mg/L) | 0.347 | 0.355 | -0.318 | 0.006** | 0.019** |
| Hemoglobin (g/L) | 0.348 | 0.358 | -0.321 | 0.069 | 0.087 |
| NL ratio | 0.351 | 0.359 | -0.323 | 0.966 | 0.840 |
| Platelet count (10<sup>9</sup>/L) | 0.335 | 0.344 | -0.308 | 0.000** | 0.000** |
| Creatinine (mg/dL) | 0.354 | 0.361 | -0.325 | 1.000 | 1.000 |
| BUN (mg/dL) | 0.347 | 0.355 | -0.320 | 0.000** | 0.001** |
| Lactate (mmol/L) | 0.348 | 0.356 | -0.320 | 0.144 | 0.459 |
| Sodium (mmol/L) | 0.352 | 0.359 | -0.324 | 0.689 | 0.957 |
| Bicarbonate (mEq/L) | 0.353 | 0.360 | -0.325 | 0.999 | 1.000 |
| pH | 0.352 | 0.360 | -0.323 | 0.805 | 0.925 |
| pO2 (mmHg) | 0.349 | 0.358 | -0.321 | 0.554 | 0.678 |
| pCO2 (mmHg) | 0.353 | 0.361 | -0.324 | 0.996 | 1.000 |

BUN: blood urea nitrogen; UBRE: Unbiased risk estimator; D1: multivariate Wald test; D2: pools test statistics from the repeated analyses; NL: neutrophils-to-lymphocytes count ratio; SF: SpO<sub>2</sub>/FiO<sub>2</sub> ratio
\*\* Variable included in final model (p-value < 0.05)

**Table S4.** L1 penalized shrunk coefficients and scaled coefficients from LASSO logistic regression.

| Variable | Coefficients | Scaled coefficients (× 3) |
| --- | --- | --- |
| Age (years) |  |  |
| < 60 | - | 0 |
| 60 - 69 | 0.413 | 1 |
| 70 - 79 | 0.935 | 3 |
| ≥ 80 | 1.666 | 5 |
| Number of comorbidities |  |  |
| ≤ 1 | - | 0 |
| > 1 | 0.353 | 1 |
| SF ratio |  |  |
| > 315.0 | - | 0 |
| 235.1 – 315.0 | 0.431 | 1 |
| 150.1 – 235.0 | 1.001 | 3 |
| ≤ 150.0 | 1.880 | 6 |
| C reactive protein (mg/L) |  |  |
| < 100 | - | 0 |
| ≥ 100 | 0.476 | 1 |
| Blood urea nitrogen (mg/dL) |  |  |
| < 42 | - | 0 |
| ≥ 42 | 0.905 | 3 |
| Platelet count (10<sup>9</sup>/L) |  |  |
| > 150 | - | 0 |
| 100 -150 | 0.288 | 1 |
| < 100 | 0.667 | 2 |
| Heart rate (bpm) |  |  |
| ≤ 90 | - | 0 |
| 91 – 130 | 0.185 | 1 |
| ≥ 131 | 0.503 | 2 |
| Intercept | -2.965 | -9 |

LASSO: least absolute shrinkage and selection operator logistic regression, SF ratio: SpO<sub>2</sub>/FiO<sub>2</sub> ratio

**Table S5.** Sensitivity analysis - Discrimination and model overall performance within complete cases.

| Model | Derivation Cohort |  | Brazilian Validation Cohort |  |
| --- | --- | --- | --- | --- |
|  | AUROC (95%CI) | Brier Score | AUROC (95%CI) | Brier Score |
| GAM | 0.871 (0.866; 0.875) | 0.108 | 0.880 (0.878; 0.887) | 0.094 |
| LASSO | 0.824 (0.792; 0.856) | 0.115 | 0.858 (0.793; 0.922) | 0.092 |
| ABC<sub>2</sub>-SPH | 0.841 (0.824; 0.858) | 0.114 | 0.852 (0.820; 0.884) | 0.107 |

GAM: generalized additive models; LASSO: least absolute shrinkage and selection operator logistic regression

**Table S6.** TRIPOD checklist for transparent reporting on a multivariable prognostic model.

| **Section/topic** | **Item** | **Checklist item** | **Page** |
| --- | --- | --- | --- |
| **Title and abstract** |  |  |  |
| Title | 1 | Identify the study as developing and/or validating a multivariable prediction model, the target population, and the outcome to be predicted | 1 |
| Abstract | 2 | Provide a summary of objectives, study design, setting, participants, sample size, predictors, outcome, statistical analysis, results, and conclusions. | 8 |
| **Introduction** |  |  |  |
| Background and objective | 3a | Explain the medical context (including whether diagnostic or prognostic) and rationale for developing or validating the multivariable prediction model, including references to existing models | 10 |
|  | 3b | Specify the objectives, including whether the study describes the development or validation of the model or both | 10-11 |
| **Methods** |  |  |  |
| Source of data | 4a | Describe the study design or source of data (e.g., randomized trial, cohort, or registry data), separately for the development and validation data sets, if applicable | 11 |
|  | 4b | Specify the key study dates, including start of accrual; end of accrual; and, if applicable, end of follow-up | 11 |
| Participants | 5a | Specify key elements of the study setting (e.g., primary care, secondary care, general population) including number and location of centres | 11 |
|  | 5b | Describe eligibility criteria for participants | 11, 12 |
|  | 5c | Give details of treatments received, if relevant | NA |
| Outcome | 6a | Clearly define the outcome that is predicted by the prediction model, including how and when assessed | 12 |
|  | 6b | Report any actions to blind assessment of the outcome to be predicted | NA |
| Predictors | 7a | Clearly define all predictors used in developing the multivariable prediction model, including how and when they were measured | 12 |
|  | 7b | Report any actions to blind assessment of predictors for the outcome and other predictors | NA |
| Sample size | 8 | Explain how the study size was arrived at | NA |

| Section/topic | Item | Checklist item | Page |
| --- | --- | --- | --- |
| Missing data | 9 | Describe how missing data were handled (e.g., complete-case analysis, single imputation, multiple imputation) with details of any imputation method | 13 |
| Statistical analysis methods | 10a | Describe how predictors were handled in the analyses | 13, 14 |
|  | 10b | Specify type of model, all model-building procedures (including any predictor selection), and method for internal validation | 13 |
|  | 10c | For validation, describe how the predictions were calculated | 14 |
|  | 10d | Specify all measures used to assess model performance and, if relevant, to compare multiple models | 14 |
|  | 10e | Describe any model updating (e.g., recalibration) arising from the validation, if done | NA |
| Risk groups | 11 | Provide details on how risk groups were created, if done | 14 |
| Development vs. validation | 12 | For validation, identify any differences from the development data in setting, eligibility criteria, outcome, and predictors | 14 |
| **Results** |  |  |  |
| Participants | 13a | Describe the flow of participants through the study, including the number of participants with and without the outcome and, if applicable, a summary of the follow-up time. A diagram may be helpful | 15, Figure 1 |
|  | 13b | Describe the characteristics of the participants (basic demographics, clinical features, available predictors), including the number of participants with missing data for predictors and outcome | 15, Table 1 |
|  | 13c | For validation, show a comparison with the development data of the distribution of important variables (demographics, predictors and outcome) | Table 1 |
| Model development | 14a | Specify the number of participants and outcome events in each analysis | Table 1 |
|  | 14b | If done, report the unadjusted association between each candidate predictor and outcome | NA |
| Model specification | 15a | Present the full prediction model to allow predictions for individuals (i.e., all regression coefficients, and model intercept or baseline survival at a given time point) | Table S4 |
|  | 15b | Explain how to use the prediction model | Page 16, Table 2 |
| Model performance | 16 | Report performance measures (with CIs) for the prediction model | Table 4, Table S5 |
| Model updating | 17 | If done, report the results from any model updating (i.e., model specification, model performance) | NA |
| **Discussion** |  |  |  |
| Limitations | 18 | Discuss any limitations of the study (such as nonrepresentative sample, few events per predictor, missing data) | 22, 23 |
| Interpretation | 19a | For validation, discuss the results with reference to performance in the development data, and any other validation data | 18-25 |
|  | 19b | Give an overall interpretation of the results, considering objectives, limitations, results from similar studies, and other relevant evidence | 18, 19 |
| Implications | 20 | Discuss the potential clinical use of the model and implications for future research | 23, 24 |
| **Other information** |  |  |  |
| Supplementary information | 21 | Provide information about the availability of supplementary resources, such as study protocol, Web calculator, and data sets | 16, 28 |
| Funding | 22 | Give the source of funding and the role of the funders for the present study | 27, 28 |

**Table S7.**
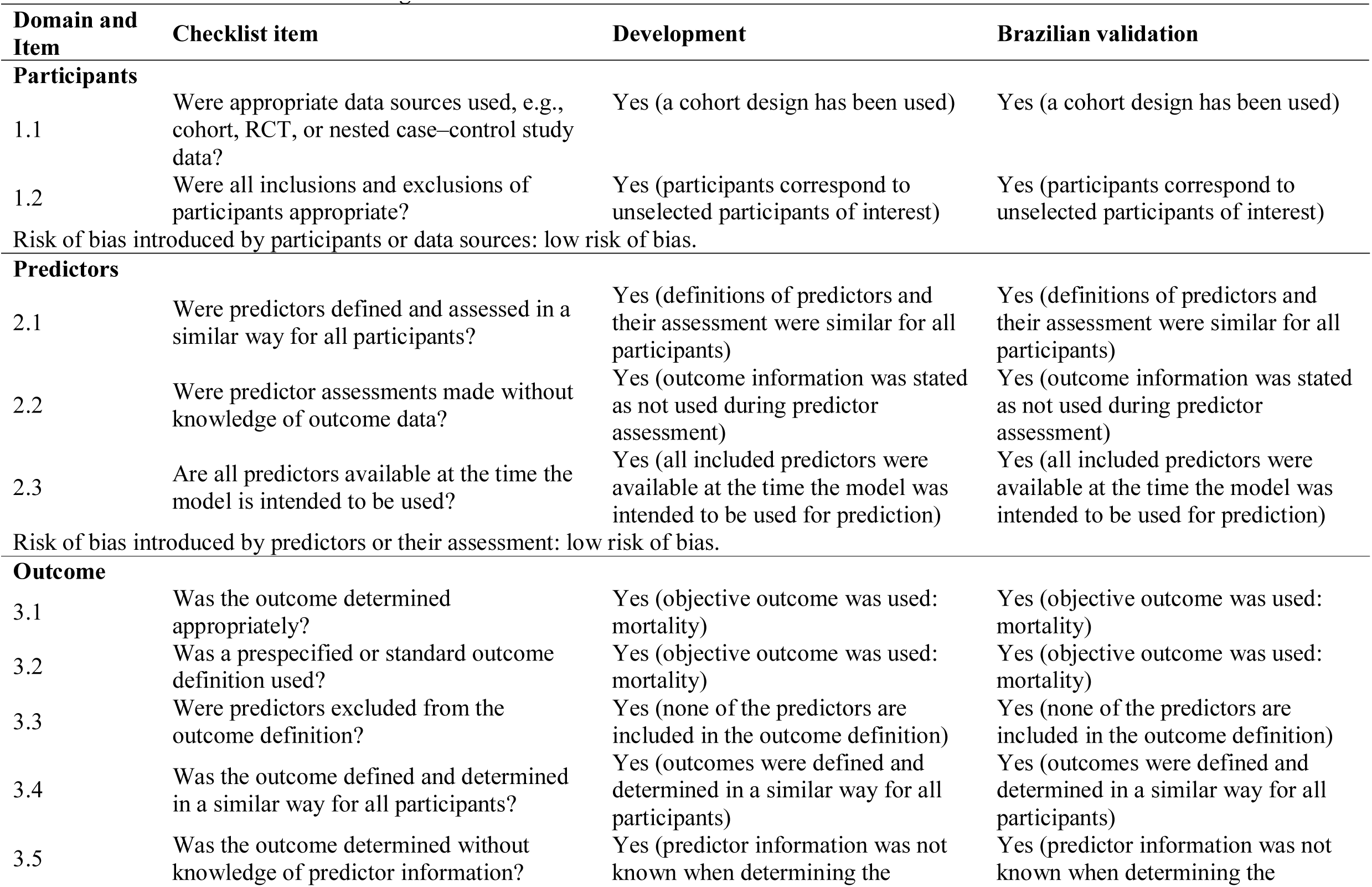
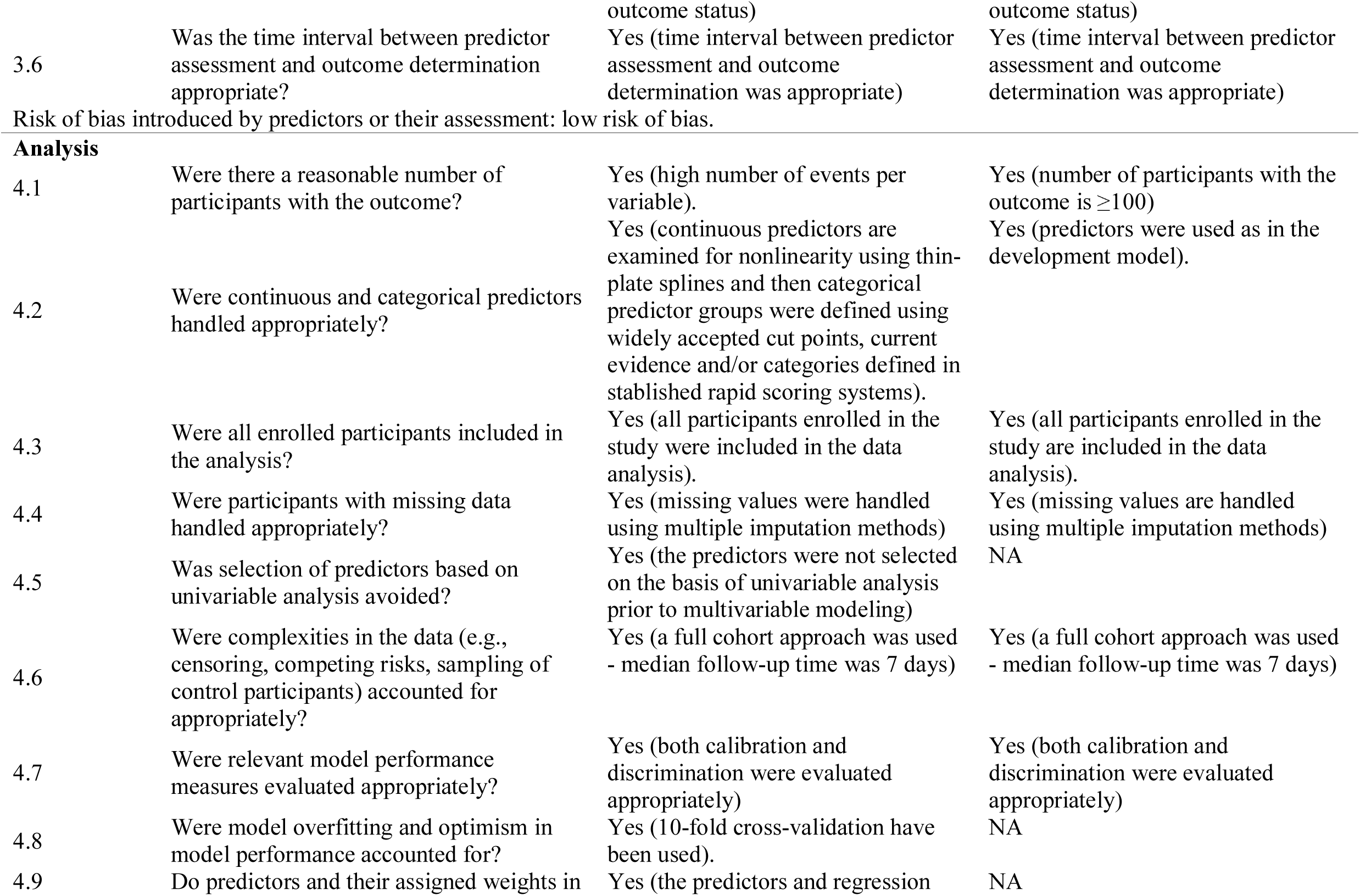
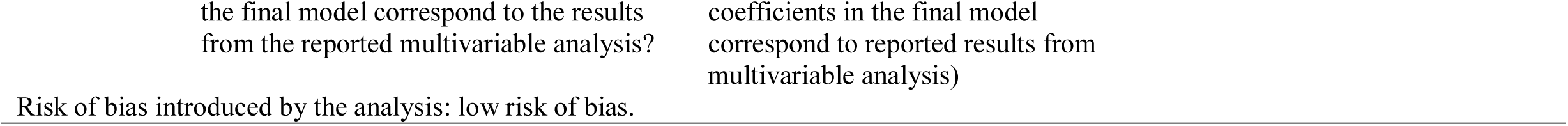
Risk of bias assessment using PROBAST checklist.

**Table S8.**
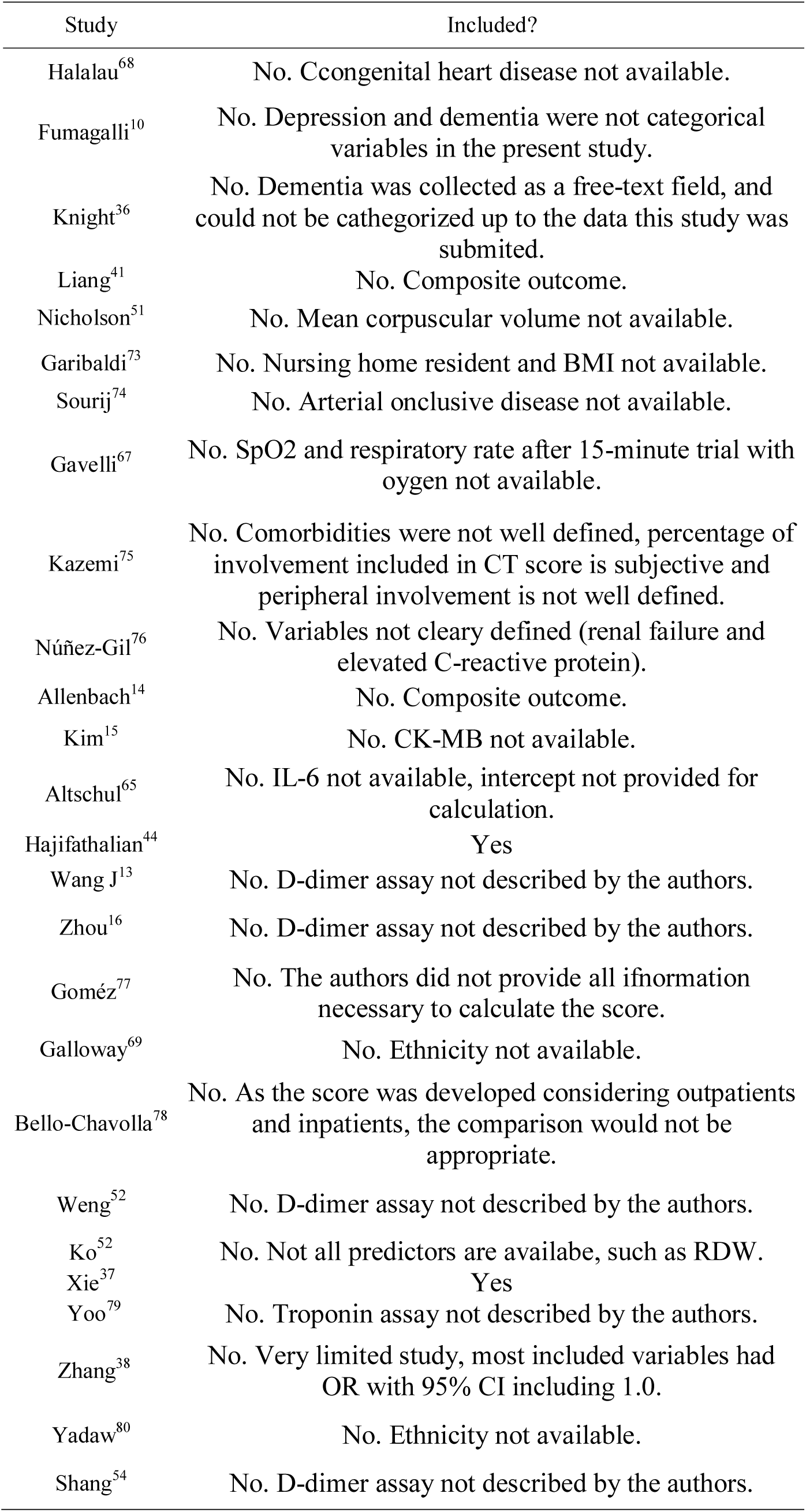
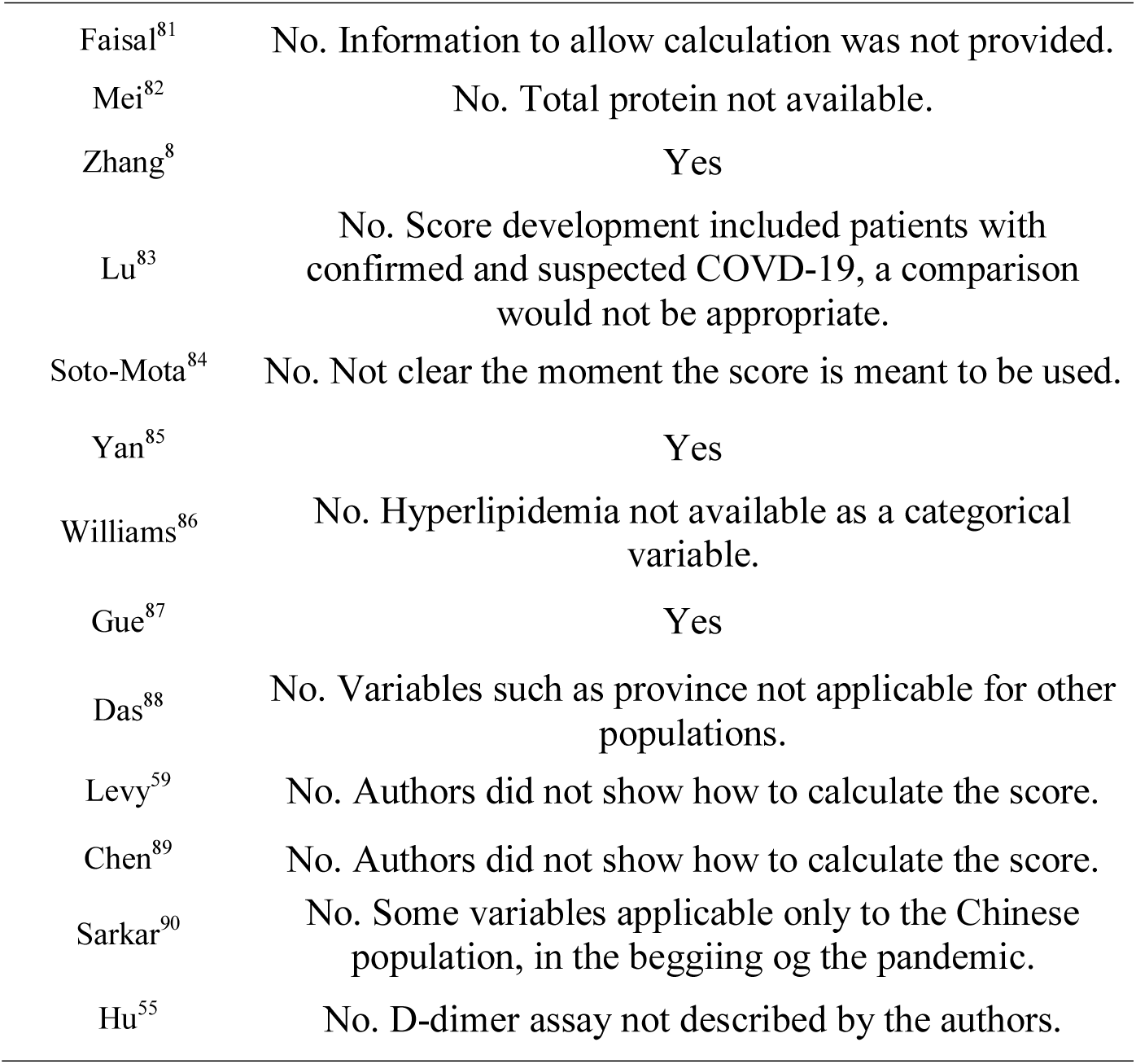
Justifications for inclusion or exclusion.

| Study | Included? |
| --- | --- |
| Halalau<sup>68</sup> | No. Congenital heart disease not available. |
| Fumagalli<sup>10</sup> | No. Depression and dementia were not categorical variables in the present study. |
| Knight<sup>36</sup> | No. Dementia was collected as a free-text field, and could not be categorized up to the data this study was submitted. |
| Liang<sup>41</sup> | No. Composite outcome. |
| Nicholson<sup>51</sup> | No. Mean corpuscular volume not available. |
| Garibaldi<sup>73</sup> | No. Nursing home resident and BMI not available. |
| Sourij<sup>74</sup> | No. Arterial occlusive disease not available. |
| Gavelli<sup>67</sup> | No. SpO2 and respiratory rate after 15-minute trial with oxygen not available. |
| Kazemi<sup>75</sup> | No. Comorbidities were not well defined, percentage of involvement included in CT score is subjective and peripheral involvement is not well defined. |
| Núñez-Gil<sup>76</sup> | No. Variables not clearly defined (renal failure and elevated C-reactive protein). |
| Allenbach<sup>14</sup> | No. Composite outcome. |
| Kim<sup>15</sup> | No. CK-MB not available. |
| Altschul<sup>65</sup> | No. IL-6 not available, intercept not provided for calculation. |
| Hajifathalian<sup>44</sup> | Yes |
| Wang J<sup>13</sup> | No. D-dimer assay not described by the authors. |
| Zhou<sup>16</sup> | No. D-dimer assay not described by the authors. |
| Gómez<sup>77</sup> | No. The authors did not provide all information necessary to calculate the score. |
| Galloway<sup>69</sup> | No. Ethnicity not available. |
| Bello-Chavolla<sup>78</sup> | No. As the score was developed considering outpatients and inpatients, the comparison would not be appropriate. |
| Weng<sup>52</sup> | No. D-dimer assay not described by the authors. |
| Ko<sup>52</sup> | No. Not all predictors are available, such as RDW. |
| Xie<sup>37</sup> | Yes |
| Yoo<sup>79</sup> | No. Troponin assay not described by the authors. |
| Zhang<sup>38</sup> | No. Very limited study, most included variables had OR with 95% CI including 1.0. |
| Yadaw<sup>80</sup> | No. Ethnicity not available. |
| Shang<sup>54</sup> | No. D-dimer assay not described by the authors. |
| Faisal<sup>81</sup> | No. Information to allow calculation was not provided. |
| Mei<sup>82</sup> | No. Total protein not available. |
| Zhang<sup>8</sup> | Yes |
| Lu<sup>83</sup> | No. Score development included patients with confirmed and suspected COVID-19, a comparison would not be appropriate. |
| Soto-Mota<sup>84</sup> | No. Not clear the moment the score is meant to be used. |
| Yan<sup>85</sup> | Yes |
| Williams<sup>86</sup> | No. Hyperlipidemia not available as a categorical variable. |
| Gue<sup>87</sup> | Yes |
| Das<sup>88</sup> | No. Variables such as province not applicable for other populations. |
| Levy<sup>59</sup> | No. Authors did not show how to calculate the score. |
| Chen<sup>89</sup> | No. Authors did not show how to calculate the score. |
| Sarkar<sup>90</sup> | No. Some variables applicable only to the Chinese population, in the beginning of the pandemic. |
| Hu<sup>55</sup> | No. D-dimer assay not described by the authors. |

**Figure S1.**
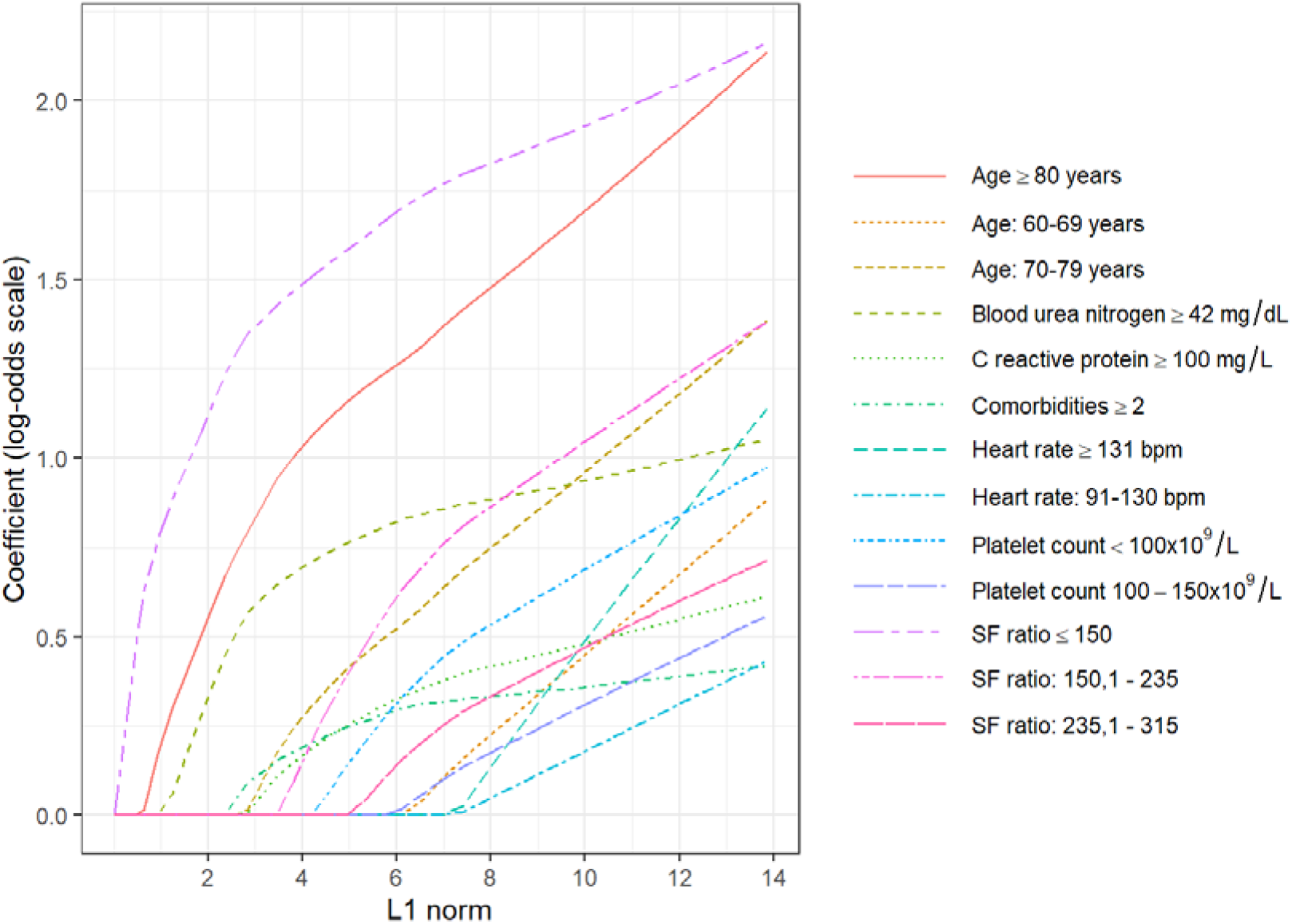
Least absolute shrinkage and selection operator logistic regression (LASSO) trace plot.

**Figure S2.**
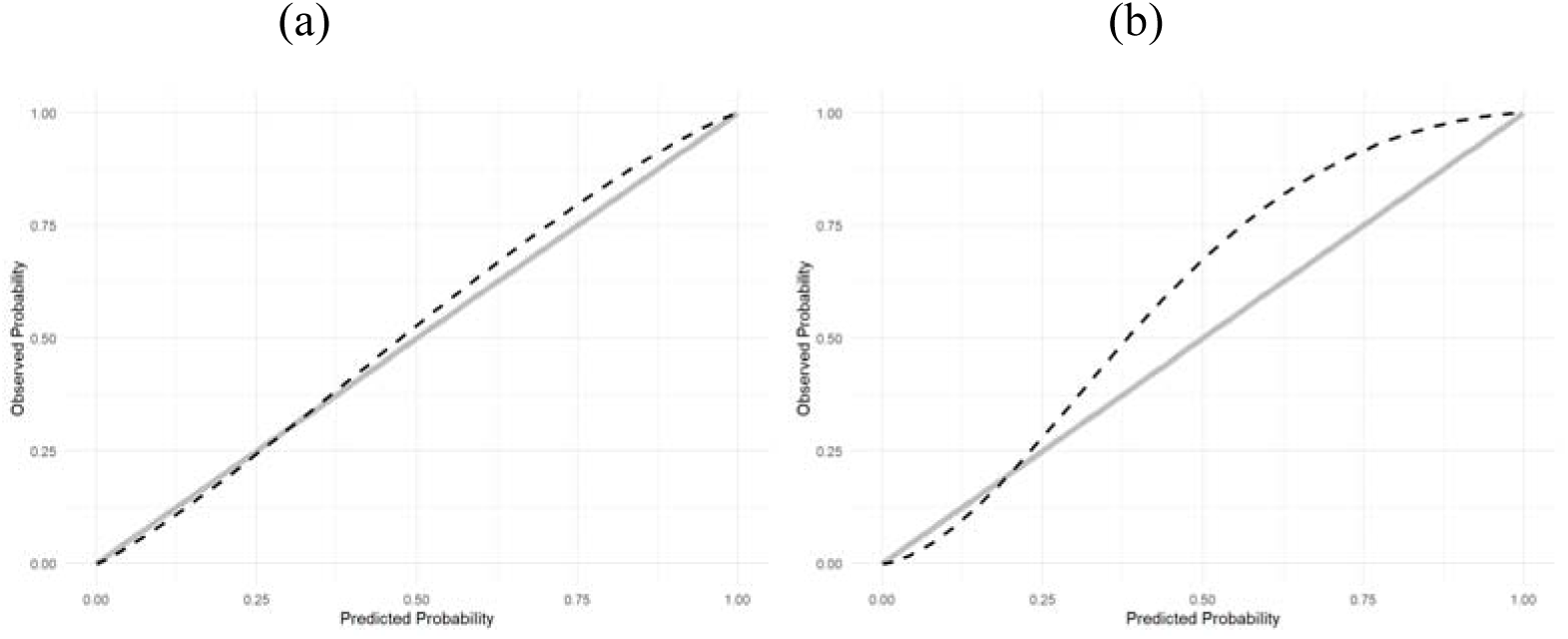
Calibration plot of ABC_2_-SPH Score in (a) Brazilian and (b) Spanish external validation cohorts.

